# Haploinsufficiency underlies the neurodevelopmental consequences of *SLC6A1*/GAT-1 variants

**DOI:** 10.1101/2022.03.09.22271804

**Authors:** Dina Buitrago Silva, Marena Trinidad, Alicia Ljungdahl, Jezrael L. Revalde, Geoffrey Y. Berguig, William Wallace, Cory S. Patrick, Lorenzo Bomba, Michelle Arkin, Shan Dong, Karol Estrada, Keino Hutchinson, Jonathan H. LeBowitz, Avner Schlessinger, Katrine M. Johannesen, Rikke S. Møller, Kathleen M. Giacomini, Steven Froelich, Stephan J. Sanders, Arthur Wuster

## Abstract

Heterozygous variants in the GAT-1 GABA transporter encoded by *SLC6A1* are associated with seizures, developmental delay, and autism. The majority of affected individuals carry missense variants, many of which are recurrent germline *de novo* mutations, raising the possibility of gain-of-function effects. To understand the functional consequences, we performed an *in vitro* GABA uptake assay for 213 unique variants, including 24 control variants. *De novo* variants consistently resulted in a decrease in GABA uptake, in keeping with haploinsufficiency underlying all neurodevelopmental phenotypes. Where present, ClinVar pathogenicity reports correlated well with GABA uptake data; the functional data can inform future reports for the remaining 72% of unscored variants. Surface expression was assessed for 86 variants; two-thirds of loss-of-function missense variants prevented GAT-1 from being present on the membrane while GAT-1 was on the surface but with reduced activity for the remaining third. Surprisingly, recurrent *de novo* missense variants showed moderate loss-of-function effects that reduced GABA uptake with no evidence for dominant negative or gain-of-function effects. Using linear regression across multiple missense severity scores to extrapolate the functional data to all potential *SLC6A1* missense variants, we observe an abundance of GAT-1 residues that are sensitive to substitution. The extent of this missense vulnerability accounts for the clinically observed missense enrichment; overlap with hypermutable CpG sites accounts for the recurrent missense variants. Strategies to increase the expression of the wildtype *SLC6A1* allele are likely to be beneficial across neurodevelopmental disorders, though the developmental stage and extent of required rescue remain unknown.

## Introduction

Large-scale exome sequencing studies have identified the gene *SLC6A1* (Solute carrier family 6 member 1, ENSG00000157103), which encodes the GABA transporter ‘GAT-1’, as a major cause of neurodevelopmental disorders. Genome-wide significant association has been reported for rare heterozygous variants in independent cohorts of developmental delay^1^, autism spectrum disorder^2^, and pediatric-onset epilepsy, particularly epilepsy with myoclonic-atonic seizures (EMAS, previously myoclonic-atonic seizures or MAE)^3^; it has also been implicated in schizophrenia^4,5^. Across these disorders, the incidence related to *SLC6A1* variants is estimated to be 2.4-2.9 in 100,000 births^6^, making it a relatively common single-gene disorder. These features make *SLC6A1*/GAT-1 a promising target for novel therapeutics.

Realizing the therapeutic potential of *SLC6A1* requires understanding the functional impact of the genetic variants observed in different phenotypes. GAT-1 is predominantly found embedded in the cell surface membrane and transports the inhibitory neurotransmitter GABA from the synaptic cleft into presynaptic neurons^7^, which may limit the inhibition of postsynaptic neurons and prepare the presynaptic neuron for further GABA release. *SLC6A1* is very highly expressed in *SV2C*/*LAMP5*-expressing GABAergic (inhibitory) neurons, highly expressed in other classes of GABAergic neurons (expressing *PVALB*, *VIP*, or *SST*), and weakly expressed in multiple non-neuronal cell types, including astrocytes, oligodendrocytes, oligodendrocyte precursor cells (OPCs), and endothelial cells^8^; there is minimal expression in excitatory glutamatergic neurons. It is widely expressed across brain regions, with expression increasing rapidly during mid to late fetal development, especially in the striatum, before reaching a steady state from birth to late adulthood^9^.

The majority of *SLC6A1* variants associated with human disorders are predicted to be missense variants or in-frame indels, some of which are recurrent *de novo* mutations (e.g., 11 individuals with p.Ala288Val, 10 individuals with p.Val342Met, **Fig. S1**); this distribution is highly suggestive of a gain-of-function mechanism. Conversely, there are also multiple individuals with protein-truncating variants (PTVs, including stop gain, frameshift, and canonical splice site variants) suggesting a co-existing loss-of-function mechanism. The function of 29 *SLC6A1* variants has previously been characterized using GABA uptake assays, including four PTVs resulting in complete loss of GABA uptake and 25 missense/in-frame variants with varying degrees of loss-of-function^10–14^ (**Table S1**). Given the loss-of-function outcomes observed, it remains unclear why *SLC6A1* is strongly enriched for missense/in-frame variants rather than the PTVs with clear loss-of-function mechanisms that are observed in most genes associated with neurodevelopmental disorders^2^. Possibilities include undiscovered gain-of-function variants, genotype-phenotype relationships that vary by disorder, as observed in *SCN2A*^15^, prenatal lethality, or a dominant negative effect, as observed in *SLC30A2*^16^.

Here, we present data on the functional impact on GABA uptake of 213 *SLC6A1* variants, 185 of which have not previously been characterized, including variants associated with schizophrenia, the majority of recurrent variants, and ‘control’ variants that are synonymous, common, or documented as benign in ClinVar. We assess the surface expression of 86 of these variants and explore the results in the context of the previously published GAT-1 structure^17^ with updated annotations of transmembrane domains, and intra- and extra-cellular loops. In addition, we explore the possibility of gain-of-function effects in stable lines for ten variants and dominant negative effects for seven variants. Finally, through integrative analysis of genomic data, we demonstrate that the GAT-1 protein is ‘delicate’ so that multiple missense variants can lead to clinically relevant haploinsufficiency. This leads to a parsimonious model of *SLC6A1*-related disorders via a heterozygous loss-of-function mechanism, with the observed missense enrichment explained by GAT-1 missense sensitivity and the recurrent missense variants explained by hypermutable CpG loci.

## Materials and Methods

Variant selection and functional assays were performed independently by teams at BioMarin and UCSF before integrating the data for a combined analysis of 213 unique variants. Combined methods are described below, with distinctions highlighted between groups as required. For detailed methods from each research group please see **supplemental methods** at the end of this manuscript.

### Variant selection

Individuals with variants in *SLC6A1* were identified from multiple sources, including ClinVar^18^, gnomAD^19^, multiple cohort studies^1–4^, case series and reports^20^, and three previously undescribed cases from the Møller group in Denmark. Across these sources, 400 unique individuals were identified in total (**Table S1**), mapping to 323 unique variants in individuals plus a further ten common missense variants from gnomAD (**Table S2**). Variants were selected for functional interpretation based on: recurrence across multiple individuals, variants in individuals with detailed phenotyping data, distribution across phenotypes, distribution across the GAT-1 protein, distribution of predicted effect on GAT-1, and distribution of population frequency. The BioMarin group assayed 181 variants, the UCSF group assayed 100 variants, and 68 of these variants were assayed by both groups (**Table S2**). Twenty-four variants were selected as controls, composed of six synonymous variants, ten missense variants observed in multiple individuals in gnomAD population controls^19^, and seven missense variants predicted to be benign from clinical sequencing as reported in ClinVar^18^. The full list of individuals and phenotypes is reported in **Table S1** and the full list of variants, genomic annotations, and functional results are reported in **Table S2**.

### Annotation of *SLC6A1* variants

Variant impact was estimated using the ENST00000287766.10 transcript for the ENSG00000157103.12 *SLC6A1* gene as defined by GENCODE v39. All reported *SLC6A1* variants in ClinVar were downloaded on June 22nd, 2022 to annotate clinical significance. All *SLC6A1* variants in the “non-neuro” version of gnomADv2.1.1 were downloaded on Aug 18th, 2022 to annotate gnomAD population allele frequency. gnomAD variant frequency estimates across all ancestries were used. Predicted missense severity was annotated using annoVar protocol ‘dbnsfp42a’ and build ‘hg38’^21^.

### *SLC6A1* gene constructs and GFP-tagging

The *SLC6A1* consensus coding sequence (CCDS: CCDS2603.1, NM_003042) was used for the creation of both gene constructs from each group under the cytomegalovirus (CMV) promoter, but with different vectors. The resulting wildtype (WT) *SLC6A1* constructs from each group were sent to Genscript (Piscataway, NJ) to perform site-specific mutagenesis to generate 181 distinct variants (BioMarin team) and 100 distinct variants (UCSF group). Genscript internally confirmed the quality and sequence of all plasmids and the expected variant was further confirmed by Sanger sequencing.

For the GFP-tagged plasmids for 86 missense variants, the 249 amino acid monomeric superfolder green fluorescent protein (sfGFP, ASSN: ASL68970) was first synthesized by Genscript. Separately, the UCSF research group sent the WT-SLC6A1 construct back to Genscript, and the sfGFP was cloned into the c-terminal end of the *SLC6A1* transcript with no spacer or linker. The final plasmid, WT-SLC6A1-sfGFP, was verified by Genscript and tested in the [^3^H]-GABA transport assay to compare relative uptake to WT-SLC6A1. No difference in GABA uptake was observed. To create the 86 GFP-tagged variant plasmids, Genscript performed site-directed mutagenesis on the WT-SLC6A1-sfGFP plasmid and verified every individual variant by Sanger sequencing.

### Preparation of cell lines and transfection of variants

Both research groups prepared human embryonic kidney cells (HEK293) separately to study the functional effects of *SLC6A1* variants. The BioMarin team employed CRISPR/Cas9 to generate a *SLC6A1* deficient HEK293T cell line (see **supplemental methods**). The GABA uptake assay was used to confirm loss of GAT-1 activity in the knockout line. The UCSF research group used the HEK293 Flp-In integration system and an empty vector (EV) control in every transfection experiment to control for any background GABA uptake in the cells (see **supplemental methods**). The [^3^H]-GABA transport assay was used before experimental design to compare background activity (EV) to wildtype activity (WT-*SLC6A1*). Plasmid sequences are reported in **Table S3**.

### GABA uptake assay and [^3^H]-GABA Transport Assay

The BioMarin research team employed Ultra-high performance liquid chromatography-MS/MS (UPLC-MS/MS) in their experimental design to functionally characterize controls and variants. Beta-lactamase (BLA) activity was measured to account for transfection variability. The UCSF research group used a radiolabeled uptake assay to characterize GABA transport of variants and controls. Both groups used the Pierce BCA Protein Assay (Thermo Fisher Scientific, Cat. 23227) to measure total protein in each well, accounting for plating variability. Refer to the **supplemental methods** for further detailing of each assay.

### Combining uptake data across cohorts

For each cohort (BioMarin and UCSF) the data from the three to six replicates (**Supplementary Table S4**) that passed quality control (count) was used to estimate the mean. Log-transformed mean estimates were compared between the two groups. Based on the strong correlation values observed between the functional data generated by BioMarin and UCSF (**Fig. S2**), we calculated the mean of GABA uptake from the 68 variants where both groups had estimated GABA uptake. We made one exception, the insertion chr3:11020264:T:TA (hg38), c.523_524insA, p.Ser175TyrFs*32 is predicted to induce a frameshift variant and premature stop codon in GAT-1. The UCSF data predicted no GABA uptake (−102.4%) while the BioMarin data predicted reduced update (−73.2%). Given that 22 other protein truncating variants had yielded a GABA uptake value below −92.3%, we elected to choose the UCSF value of −102.4% for the combined result.

### Defining thresholds for functional impact

The GABA uptake values for 24 control variants (Table S1) were normally distributed around a mean of −0.8% with a standard deviation of 24.7%. We used z-score thresholds of 1.96 standard deviations (48.4%, p≤0.05, two-sided) to define loss-of-function (−0.8% − 48.4% = −49.2%) and gain-of-function (−0.8% + 48.4% = 47.5%) and 3.29 standard deviations (81.2%, p≤0.001, two-sided) to define severe loss-of-function (−0.8% − 81.2% = −82.1%). No variants were above 80.4%, which would be the equivalent severe gain-of-function threshold.

### Immunostaining

For immunostaining experiments, HEK293 cells were plated on poly-D-lysine treated 96-well black PhenoPlate (Perkin Elmer Health Sciences, Inc) with an optically clear flat-bottom and at an optimized cell density of 2.4×10^4^ cells/well. All wells on the edges of the plate were excluded to prevent edge effects. Cells were reverse transfected with respective SLC6A1-sfGFP-mutant plasmid and controls (WT-SLC6A1-sfGFP and EV-sfGFP). Mutant plasmids were transfected in 3 biological replicates and performed in duplicate. WT and EV were treated the same but had an additional plate with 12 biological replicates per plasmid and performed in duplicate. After 48 hours, cells were washed once with Hank’s Balanced Salt Solution (HBSS) and then the plasma membrane was stained with Wheat Germ Agglutin Alexa Fluor 647 conjugate (Invitrogen Life Sciences Corporation) diluted (1:1000) in cold HBSS for 5 minutes at room temperature. The stain was then aspirated and cells were carefully washed three times with HBSS. Cells were fixed with 3.7% formaldehyde in HBSS for 15 minutes. The fixing solution was then aspirated and cells were washed again three times with HBSS. Next, the cytoplasm was stained using HCS CellMask Orange Stain (Invitrogen Life Sciences Corporation) in HBSS (1:1000) for 5 minutes at room temperature and kept in darkness. The stain was aspirated and cells were washed two more times with HBSS. Lastly, the nucleus was stained with Hoescht in HBSS (1:2000) for 20 minutes at room temperature and kept in darkness. Cells were washed twice with HBSS and 100 µL of buffer was left in the wells for imaging. Imaging was performed within 24 hours of staining or the same day and then stored in a 4°C fridge wrapped in aluminum foil.

### High-throughput confocal imaging with IN Cell 6500HS Analyzer and IN Carta Software analysis

Immunostained plates were imaged with the IN Cell Analyzer 6500HS (GE Healthcare, WA, USA). All images were taken with a Nikon 40x objective with the laser and software autofocus (AF) applied. A total of eight fields of view were captured per well across all plates (n=48 total per plasmid: eight fields of view, three biological replicates, performed twice). Four channels were used to capture the respective stain or fluorescence. The blue laser (405 nM excitation and 455/50 nM emission) captured the Hoechst stain. The orange laser (561 nM excitation and 605/51 nM emission) captured the orange HCS CellMask stain. The far-red laser (642 nM excitation and 682/59 nM emission) captured the Alexa Fluor 647 conjugate. The green laser (488 nM excitation and 524/48 nM emission) captured GFP expression.

The resulting images were visualized and analyzed with the IN Carta image analysis software (Molecular Devices, LLC., San Jose, CA). The protocol segmented the images by cell components. The blue wavelength was used to segment the mono-nucleated cells. The orange wavelength was used to segment the whole cell using the orange HCS CellMask. Once the software identified the nucleus and whole cell, the membrane was segmented using the red wavelength. For example images, see **Fig. S3**. The protocol was run and Pearson correlation values were generated for GFP expression colocalization with the plasma membrane stain (Alex Fluor 647). Pearson correlation values are calculated through the software using total intensities (total intensity = intensity x cell area) of the respective wavelengths captured in the field of view(s). After all values were generated from this protocol, the Classifier Tool within the IN Carta software was applied to further refine Pearson correlation raw scores. This tool allows you to define your upper and lower bounds for intensities detected across the entire plate, helping to account for plate-to-plate variability. Lower bounds were classified as ‘low GFP’ expression or background that is detected by the software. Upper bounds were classified as an ‘overexposed GFP’ signal, which is bright puncta detected. Between the upper and lower bounds, there was a ‘High GFP’ signal and the Classifier tool reran the protocol with these newly defined classes. Newly generated values consider Pearson correlation scores by class. All final analyses in this study were conducted based on the ‘High GFP’ class (**Fig. S4**).

### Protein structure mapping in 2D and 3D (combined datasets)

To visualize the topology of *SLC6A1/*GAT-1 and percent-wildtype uptake activity of missense variants, the UCSF transmembrane protein display tool TOPO2 was used (http://www.sacs.ucsf.edu/cgi-bin/open-topo2.py). The topological domains of *SLC6A1* were annotated in two different ways (**Tables S1** and **S2**). The initial annotations were taken from UniProt (ID P30532). The second annotations, used for our primary analysis, were graciously provided by Dr. Cornelius Gati, whose group recently published the 3D structure of GAT-1^17^. All final topology figures and subsequent 3D representations were taken from the solved GAT-1 structure annotations provided by Dr. Gati. Molecular docking was performed using Glide^25^ from the Schrödinger suite. GABA was docked to the cryo-EM structure of GAT-1 (PDB: 7SK2). The ligand binding site was defined based on the coordinates of Tiagabine in the published structure. The docking results, figures, and videos were visualized via PyMOL, version 2.5.

### Data analysis and figures

Data were analyzed using Python (including pandas, NumPy, statsmodels, matplotlib, and seaborn libraries). Figures were composed using Adobe Illustrator. Further details on individual analysis can be found under **supplemental methods**.

## Results

### Impact of 213 variants in *SLC6A1* on GABA uptake activity

A combined search of publications and large-scale genomic cohorts identified 400 individuals with variants in *SLC6A1*, which mapped to 323 unique variants^4,10,11,19,20,26–33^ (**Table S1**). Our two groups (BioMarin, UCSF) independently selected subsets of these variants for functional analysis of GABA uptake using a [^3^H]-GABA transport assay in HEK293 cells (**supplemental materials and methods**). Assays of GABA uptake are reported as the percent difference from wildtype, with 0% being the same as wildtype and −100% being the complete absence of GABA uptake. In total, 213 variants in *SLC6A1* were assessed and the 68 variants analyzed by both groups yielded consistent results (R^2^=0.79, P=5.3 x 10^-24^, **Fig. S2**). Our data were also consistent with previously published results (n=21; R^2^=0.51 [^12^], n=7; R^2^=0.67 [^11^], **Fig. S2**).

The 213 variants included 24 control variants predicted not to contribute substantial risk for neurodevelopmental disorders: six synonymous variants, ten missense variants selected because they were observed in gnomAD population controls^19^, and seven missense variants predicted to be benign from clinical sequencing reported in ClinVar^18^. Across these 24 control variants, GABA uptake was equivalent to the reference (non-variant) version of *SLC6A1* (Mean=-0.8%, StDev=24.7%, Range=-46.6% to 51.0%). We used these variants to define thresholds based on z-scores for severe loss-of-function (≤-82.1%, p≤0.001, 100 variants), loss-of-function (>-82.1% and ≤-49.2%, p≤0.05, 25 variants), and gain-of-function (>47.5%, p≤0.05, 3 variants), with 85 variants in the typical range (>-49.2% to ≤47.5%) (**Fig. 1B**). These four variant effect types are highlighted on a 2D-topology representation of GAT-1 for missense variants (**Fig. 1A**). To see 2D-topology of other previously published groups (Mermer, Mattison), refer to **Fig. S5**.

**Figure 1.**
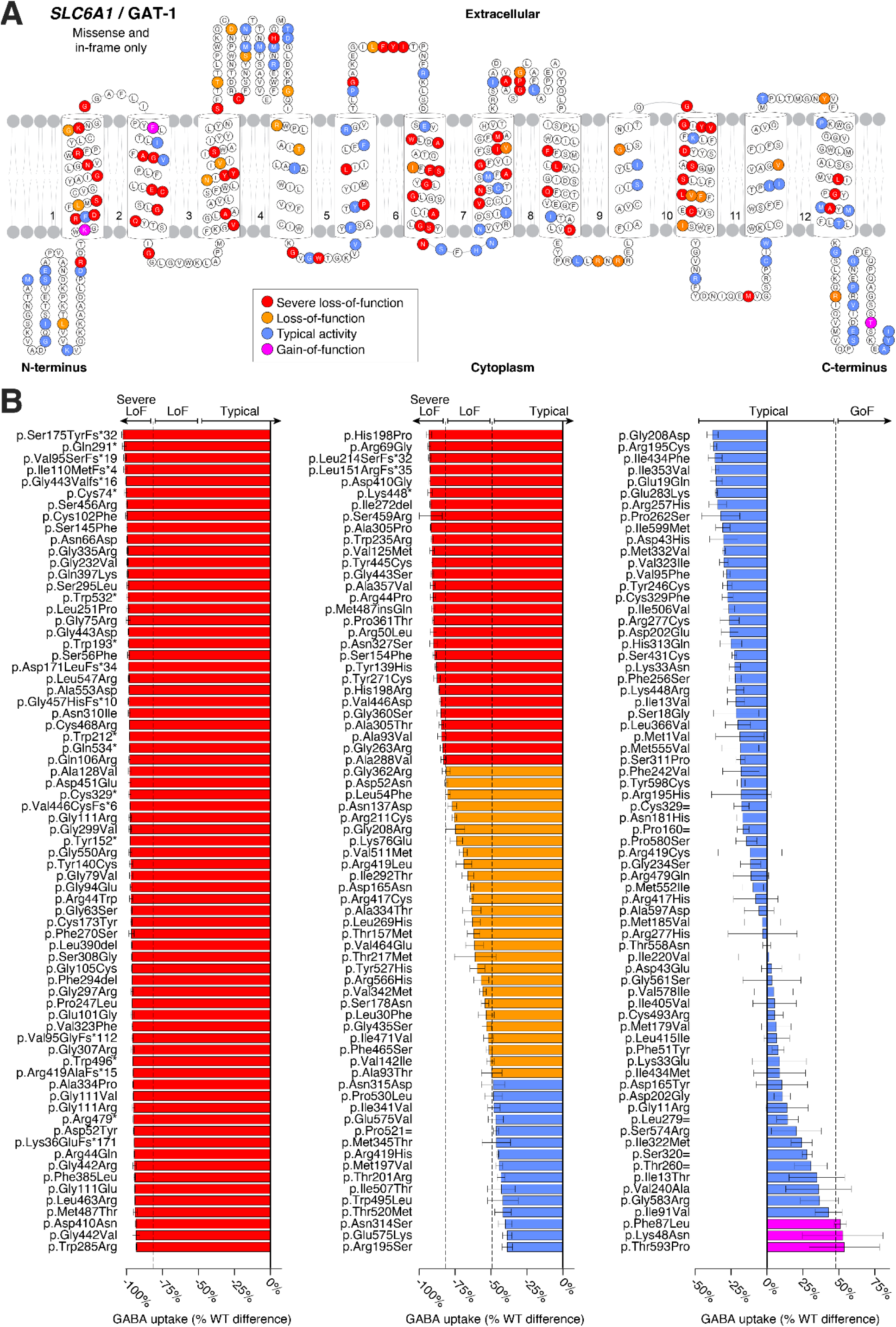
Topology of the GAT-1 protein and GABA uptake values by functional type. **A)** 2D representation of the GAT-1 protein, organized by 12 transmembrane domains and linking or terminal chains. Only missense and in-frame variants are highlighted. **B)** GABA uptake functional data as a percentage of wildtype and highlighted by activity type for 213 variants tested across the 599 amino acids of the GAT-1 protein. Individual data bars represent mean ±SEM of three biological replicates performed in triplicates. Abbreviations: GoF: gain-of-function, LoF: loss-of-function, WT: Wildtype.

### Limited evidence for gain-of-function variants

Three variants exceeded our gain-of-function threshold of >47.5% (**Fig. 1**). One of these, p.Phe87Leu, was a control variant selected from population controls in gnomAD. Another, p.Lys48Asn, was a variant of unknown significance and unknown inheritance that is also present in gnomAD that was identified in an individual with early onset epileptic encephalopathy^33^. The third one, p.Thr593Pro, was a variant of unknown significance absent from gnomAD that was reported in ClinVar without clinical details. Given the caveats of overexpressing GAT-1 in HEK293 cells for uptake assays, we sought to assess whether these represented genuine gain-of-function effects. Stable cell lines were generated for the ten variants with the highest GABA uptake values in the UCSF cohort, including p.Phe87Leu and p.Thr593Pro, but not p.Lys48Asn. These ten stable cell lines were retested for GABA uptake activity (**Fig. S6**). Across all ten, the uptake values were substantially lower than those observed in the transiently transfected cell lines and exhibited a non-significant trend to being above the wildtype levels (i.e., 100%). If variants do increase GABA uptake, the effects are small and unlikely to contribute substantially to neurodevelopmental phenotypes.

### Neurodevelopmental phenotypes are consistently associated with loss-of-function effects

All 23 protein truncating variants (PTVs) assessed resulted in almost no GABA uptake (i.e., severe loss-of-function), consistent with nonsense-mediated decay or non-functional GAT-1 protein (**Fig. 2A**). Of 166 clinically-ascertained missense variants/in-frame indels, 77 were severe loss-of-function, 27 were loss-of-function, 2 were gain-of-function, and 60 were in the typical range. Even missense variants/in-frame indels in the typical range had a median GABA uptake of −25.7%, suggesting that a subset of these may also contribute some neurodevelopmental risk via mild loss-of-function (**Fig. 2A**). Across all variants assayed, GABA uptake was correlated with population allele frequency (R^2^=0.38, P=8.4 x 10^-24^, **Fig. S7**). Of the 156 variants absent in 114,704 population controls, 115 (73.7%) resulted in severe loss-of-function or loss-of-function. In contrast, none of the variants observed in ten or more individuals showed evidence of reduced GABA uptake (**Fig. 2B**).

**Figure 2.**
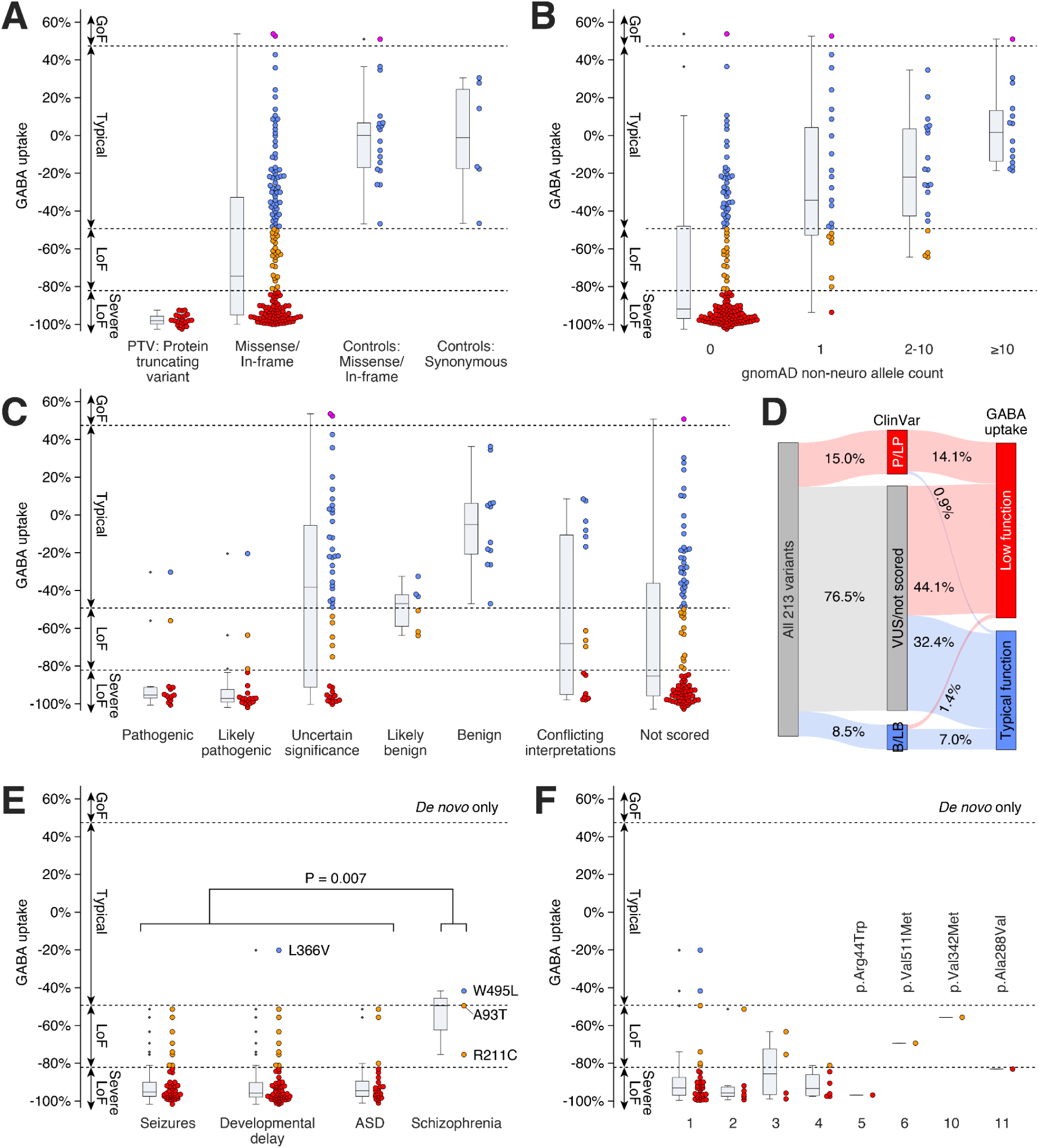
GABA uptake of *de novo* variants by phenotype and recurrence. **A)** GABA uptake data for 213 variants by predicted impact on the GAT-1 protein. Protein truncating variants and missense/in-frame variants are ascertained from clinical populations (left two categories) while 24 variants selected as controls were split between missense/in-frame and synonymous variants (right two categories) and used to set thresholds for loss-of gain-of-function (dashed lines). **B)** GABA uptake values by population allele count in 114,704 non-neuropsychiatric samples in gnomAD and **C)** by ClinVar clinical significance. **C)** A Sankey plot summarizing the proportion of variants where ClinVar category could be reclassified. **E)** GABA uptake values for *de novo* variants by the presence of seizures, developmental delay, autism spectrum disorder (ASD), or schizophrenia. If phenotypes are comorbid (e.g., seizures and developmental delay) the variant is shown for all phenotypes for which it is reported. **F)** GABA uptake values of *de novo* variants observed in single individuals (1) or multiple individuals (2-11). Each variant is shown once. The four most recurrent variants (5, 6, 10, and 11) are labeled. Abbreviations: GoF: gain-of-function, LoF: loss-of-function, A93T: p.Ala93Thr, R211C: p.Arg211Cys, W495L: p.Trp495Leu. Statistical tests: E: two-sided Wilcoxon test.

Clinical significance has previously been stated for 113 variants (**Tables S2, S5**). Where clear determinations were made, these were generally supported by the functional assay: 32 (5 PTVs, 27 missense) were reported as Pathogenic or Likely Pathogenic and 30 (93.8%) of these were indeed found to be loss-of-function, while 18 missense variants were reported as Benign or Likely Benign of which 15 (83.3%) were in the typical range (**Fig. 2C**). The remaining 163 variants had Uncertain (n=46), Conflicting (n=16), or absent (n=101) clinical significance; 94 (57.7%) of these resulted in severe loss-of-function or loss-of-function. Thus, our functional data is consistent with 97 (45.5%) of the remaining variants being clinically relevant and 71 (33.3%) variants having minimal impact on function (**Fig. 2D**).

To minimize the ascertainment bias, we assessed genotype-phenotype correlations focusing on the 67 *de novo* variants (**Fig. 2E, S8**). All but one of these variants were identified in an individual diagnosed with a neurodevelopmental disorder, the exception being p.Leu251Pro, reported in an individual with no seizures at 21 months of age and no data reported regarding autism spectrum disorder (ASD) or developmental delay (DD)^20^. The completeness of the phenotyping data varied by variant (**Table S1, S2**). Seizures were reported in 46 of the 55 (83.6%) with data, ASD reported in 24 of 44 (54.5%), and DD in 58 of 59 (98.3%). Variants were consistently loss-of-function in cases with seizures, ASD, and DD (median GABA uptake −0.95, −0.95, −0.96 respectively). The most frequently reported seizure type was Epilepsy with Myoclonic Atonic Seizures (EMAS), reported in 22 of the 37 (59.5%) for whom seizure type was available. In addition, early onset absence epilepsy (EOAE), childhood absence epilepsy (CAE), developmental and epileptic encephalopathy (DEE), and Lennox-Gastaut Syndrome (LGS) were related to reduced GABA uptake. In contrast, non-familial non-acquired focal epilepsy (NAFE) consistently resulted in typical levels of GABA uptake (**Fig. S8**), suggesting *SLC6A1* disruption is probably not associated with NAFE. No clear patterns were observed between variant effect and age of seizure onset (**Fig. S9**).

### Some variants observed in schizophrenia have loss-of-function effects

Two schizophrenia cohorts contributed *SLC6A1* missense variants that were predicted to be damaging (MPC score ≥ 2)^34^. The first focused on *de novo* variants from 3,444 cases and reported that *SLC6A1* was associated with schizophrenia (P=7.9 x 10^−5^, uncorrected)^4^. Two of the three *de novo* variants resulted in loss-of-function effects (p.Ala93Thr, p.Arg211Cys), while one was in the typical range (p.Trp495Leu). The median GABA uptake across these variants is −49.5% - a value in the loss-of-function range but higher than that observed for the neurodevelopmental phenotypes (P=0.007, two-sided Wilcoxon Test, **Fig. 2E**). The second cohort was a case-control analysis of 24,248 cases (including the previous 3,444); they reported enrichment for *SLC6A1* variants that did not meet genome-wide correction (P=0.006, uncorrected; P=0.50, corrected)^5^. While these additional variants were not reported to be *de novo* in the schizophrenia cases, two of the missense variants from this cohort were at the same location as *de novo* variants identified in neurodevelopmental disorders: p.Ala305Thr (severe loss-of-function) and p.Ala334Thr (loss-of-function). Of the remaining 11 variants in the second cohort, two resulted in severe loss-of-function (p.Asp410Asn, p.Gly442Arg), two in loss-of-function (p.Asn137Asp, p.Arg417Cys), and seven were in the typical range (**Table S2, Fig. S8**).

### Correlation of functional data and surface expression of 86 missense variants

Given the role of loss-of-function effects for the clinically relevant *SLC6A1*/GAT-1 missense variants, we sought to understand the relationship of protein trafficking mechanisms to these functional outcomes. A reduction in GAT-1 stability could lead to a relative deficit in surface colocalization^10,12^. Surface expression was assessed for 86 missense variants using a high-content imaging approach to assess the relative co-localization of GFP-tagged GAT-1 with a membrane stain. While wildtype GAT-1 is located on the plasma membrane, it is also distributed at lower levels throughout the cell body in HEK293 cells (**Fig. S4**). Given this mixed localization, the high-content imaging experimental design utilized segmentation as part of the image analysis to quantify the colocalization of GFP-tag and plasma membrane stain through the Pearson correlation colocalization (PCC) metric. The wildtype GAT-1 GFP-tagged plasmid (WT) yielded a PCC value of 0.32 (n=26), which we defined as 100% wildtype surface expression. As a negative control, we used an ‘empty vector’ (EV) plasmid that contained GFP but not GAT-1; the resulting GFP is expressed non-specifically throughout the cell, with only a small amount on the membrane (**Fig. S4**). The EV plasmid resulted in a PCC value of 0.15 (n=26 replicates), 46.9% below the wildtype GAT-1 value (**Fig. S10**). For each of the 86 missense variants, we tested six replicates (mean of 5.9 replicates after data cleaning), from which we calculated both the PCC and the percentage wildtype surface expression (**Fig. S10**). For further analysis, the percentage wildtype surface expression values below that of EV (46.9%) were increased to 46.9%.

GABA uptake data was correlated with surface expression results (R^2^=0.23, P=3×10^-6^, **Fig. 3A**). Variants with typical GABA uptake generally had high surface expression, while loss-of-function variants had highly variable surface expression. To interpret these relationships, we applied a K-means algorithm to define three groups of variants (K=3, **Fig. 3A**): Group 1) variants present on the surface with typical uptake (n=30), Group 2) variants absent from the surface with low uptake (n=40), and Group 3) variants that were present on the surface but had low uptake (n=16). Four variants with typical GABA uptake were outliers with very low surface expression: p.Gly561Ser (PCC=0.11), p.Glu283Lys (PCC=0.12), p.Pro530Leu (PCC=0.06), identified in a large-scale epilepsy sequencing project^27^ and p.Trp495Leu (PCC=0.09), a *de novo* variant identified in schizophrenia^4^ (**Fig. S11**). In groups 1 and 2, a substantial fraction of the variability in GABA uptake can be explained by relative surface expression (R^2^=0.69, P=8×10^-19^) but a different mechanism is required to explain the loss-of-function effects of the 16 missense variants in group 3.

**Figure 3.**
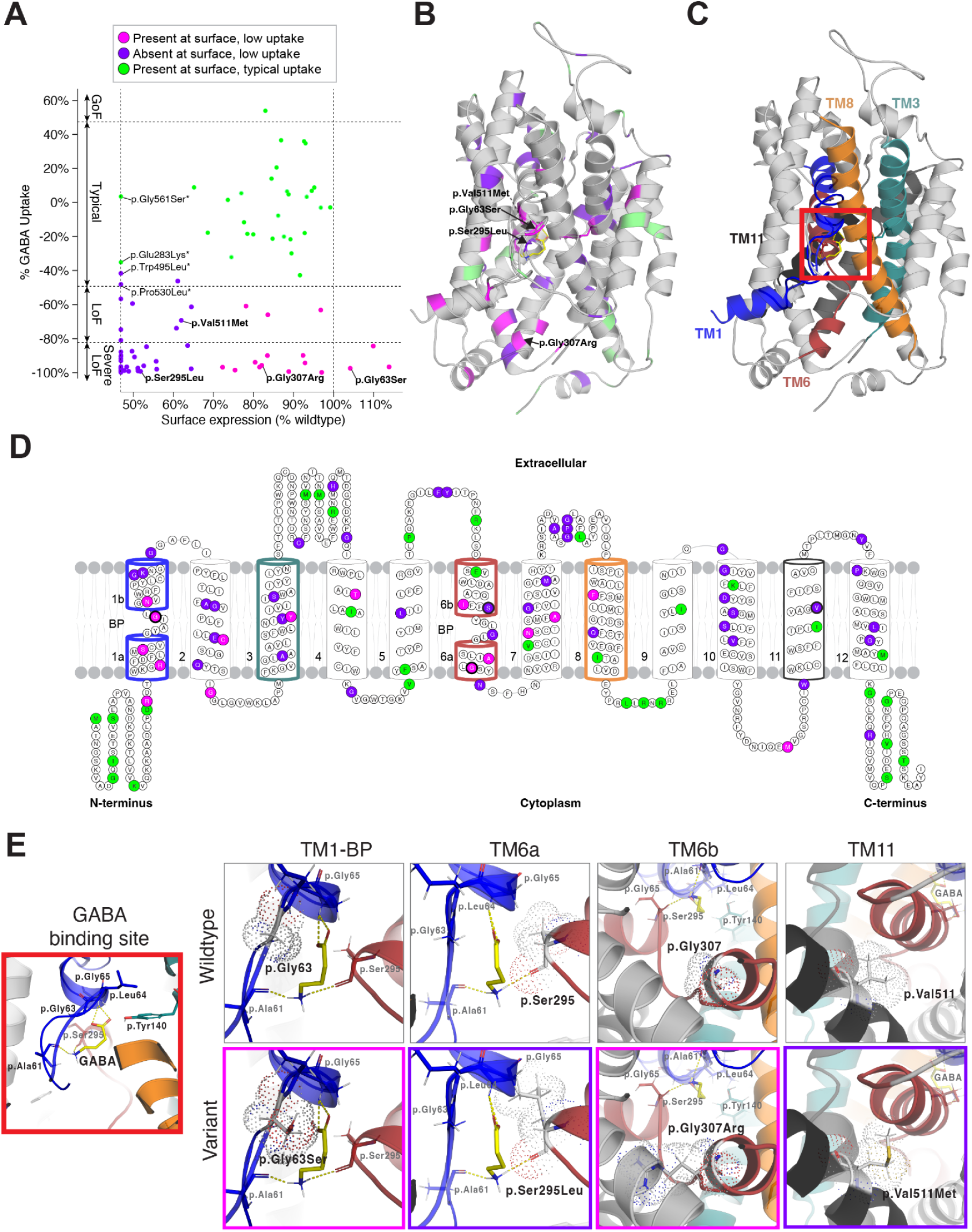
Clustering of surface expression and GABA uptake results of 86 missense variants and highlighted variants in GAT-1 structure and binding site. **A)** Individual GABA uptake values (y-axis) correlated to surface expression results (x-axis) and classified by K-means clustering as either present at the surface and low uptake (magenta), absent at the surface and low uptake (purple), or present at the surface and typical uptake (green). Variants named with an asterisk denote outliers. All other called variants are further illustrated in B and E. **B)** GAT-1 3D structure in its inward-open conformation with highlighted variants by cluster. **C)** GAT-1 3D structure repeated and highlighted by transmembrane domains. The red square indicates the binding site. **D)** 2D topology of GAT-1 structure showing individual variants colored by cluster and corresponding transmembrane domains highlighted from panel C. **E)** The GAT-1 binding site inset from panel C (red square) and relevant amino acids is shown with GABA bound. The wildtype and variant 3D structures of four variants are shown and the variant insets are boxed in colors relevant to their clustering from panel A (magenta or purple). Two variants form part of the binding site for GABA (p.Gly63Ser, p.Ser295Leu), one variant is part of TM6 (p.Gly307Arg) and the variant in TM11 is the top recurrent variant (p.Val511Met, n=6) with available surface expression data. Abbreviations: GoF: gain-of-function, LoF: loss-of-function, TM1: transmembrane domain 1, TM3: transmembrane domain 3, TM6: transmembrane domain 6, TM8: transmembrane domain 8, TM11: transmembrane domain 11

### Mapping of 86 variants to the human GAT-1 protein structure

The human GAT-1 transporter is a 599 amino acid protein that is a member of the solute carrier 6 (SLC6) transporter family, also known as sodium and chloride (Na^+^/Cl^-^) coupled transporter proteins. The SLC6 family contains 20 transporter members that are further divided into four subgroups based on substrate selectivity and sequence homology. This includes the neurotransmitter transporters *SLC6A3* (DAT; Dopamine transporter) and *SLC6A4* (SERT: Serotonin transporter), which share about ∼50% sequence identity to GAT-1. Like all other SLC6 family members, GAT-1 shares the conserved 12 transmembrane (TM) domain conformation with amino (N-) and carboxyl (C-) termini located intracellularly^35^. Previous studies have mapped out important GAT-1 substrate binding site components using the leucine transporter LeuT as a structural model, a bacterial ortholog of the eukaryotic Na^+^/Cl^-^ transporters^22,36^. From this model, the main four TM domains that form the binding pocket, or cylindrical ring, of GAT-1 are identified as TM1, TM3, TM6, and TM8. The most conserved regions across the Na^+^/Cl^-^ transporter family include TM1 and TM6, reflecting their importance in binding substrates and inhibitors^22^. Both TM1 and TM6 helices have unwound ‘hinge regions’ that separate the helices, causing a split of these regions, with the two halves labeled TM1a/TM1b, and TM6a/TM6b (**Fig. 3D**). These hinge regions are the formal binding site for GAT-1 substrates and its co-transported ions sodium (2Na^+^) and chloride (Cl^-^). More recently, the cryo-EM structure of wild-type human GAT-1 was determined in the inward-open conformation, giving additional insight into specific binding modalities and GAT-1 mechanics^17^. We mapped all 86 variants to the cryo-EM-derived structure (PDB ID: 7SK2) and closely analyzed structure-function relationships to determine if this could provide insight into the variants we identified in Group 3 (**Fig. 3B, 3C, 3D**).

Considering the 3D structure (**Fig. 3B, 3C**), we visually calculated the number of variants located on the outer surface of GAT-1 versus the inner surface for each Group^37^ (**Fig. S12, S13**). We distinguished that 80% of Group 2 variants (low uptake and poor protein trafficking) and 93.8% of Group 3 variants (low uptake, but proper protein trafficking) were found buried in the interior of the 3D structure, in contrast to only 36.4% of variants from Group 1 (typical uptake and proper protein trafficking). This may suggest that variants positioned within the internal structure can have variable impact on protein trafficking but are more likely to lead to reduced function of GAT-1. Whereas, variants that fall on the outer surface of the protein structure, and more notably the unwound intracellular and extracellular loops (Group 1, **Fig. 3D, Fig. S12A**), are less likely to cause deficits in protein trafficking or GABA uptake. Additionally, of the 16 variants in Group 3, 13 were located within transmembrane domains and eight of these are found in the transmembrane domains most closely associated with the GABA binding pocket of GAT-1 (TM1, TM3, TM6, and TM8, **Fig. 3D**), strongly suggesting that their loss-of-function effects were through disruption of this binding modality rather than protein trafficking to the membrane.

To assess this hypothesis, variants within the transmembrane domains that form the binding pocket and main intracellular gate (TM1 and TM6, p.Gly63Ser, p.Ser295Leu, p.Gly307Arg) were analyzed in more detail considering the 3D structure (**Fig. 3E**). Variant p.Gly63Ser (Group 3), a recurrent *de novo* variant seen in two individuals with seizures, severe developmental delay, and ASD, forms part of the TM1 GABA binding site (**Fig. 3E**). The wildtype residue (p.Gly63) is predicted to form a hydrogen bond with the carboxylic acid of GABA^17^. When mutated to p.Gly63Ser, a polar functional group is added, increasing strain on the binding site, which may interfere with the native hydrogen bonds to GABA and reduce uptake. Variant p.Ser295Leu (Group 2) also forms part of the GABA binding site, but within TM6. In its wildtype form, p.Ser295 forms hydrogen bonds with the backbone of GABA at the amine (−NH_2_). The polar to nonpolar p.Ser295Leu mutation is likely to weaken this GABA and/or ion binding. Furthermore, the change to the longer functional group in leucine likely causes substantial strain that interferes with the TM1 helix (**Fig. 3E**), potentially impairing protein trafficking and disrupting the opening and closing mechanics of the transporter necessary for GABA uptake. This same phenomenon can be seen in the variant p.Gly307Arg (Group 3), where changing glycine with the positively charged side chain from arginine creates a large strain and interference between TM6 (red) and TM2 (gray).

Additionally, we examined the most recurrent missense variant that was available within these 86 variants (p.Val511Met, Group 2, n=6, **Table 1**) to investigate whether its position outside of the binding pocket (TM11) might suggest an additional role underlying the observed recurrence. Interestingly, the addition of a sulfur group gives rise to one of the most hydrophobic amino acids, methionine, and is positioned within TM11. Logically, we would not expect such a substitution, where both wildtype and mutant have amino acids with hydrophobic side chains, to impact protein trafficking as we observed. Hydrophobic amino acids are known to interact with hydrophobic ligands, such as lipids or the plasma membrane^38^. No clear explanation for recurrence was observed from this analysis.

**Table 1.** Recurrent *de novo* mutations in *SLC6A1/*GAT-1.

| SLC6A1 variant | GAT-1 variant | Count | Seizures | EMAS | Absence | Mean seizure onset | Developmental delay | ASD | Schizophrenia | De novo | Inherited | GABA uptake | Mutability |
| --- | --- | --- | --- | --- | --- | --- | --- | --- | --- | --- | --- | --- | --- |
| c.863C>T | p.Ala288Val | 11 | Yes | Yes | Yes | 25mths | Yes | Yes | . | Yes | Yes | -83.0% | 0.04 |
| c.1024G>A | p.Val342Met | 10 | Yes | Yes | Yes | 28mths | Yes | Yes | . | Yes | Yes | -55.7% | 0.04 |
| c.1531G>A | p.Val511Met | 6 | Yes | . | Yes | 12mths | Yes | . | . | Yes | Yes | -69.4% | 0.04 |
| c.130C>T | p.Arg44Trp | 5 | Yes | Yes | Yes | 28mths | Yes | . | . | Yes | . | -96.8% | 0.06 |
| c.331G>A | p.Gly111Arg | 4 | Yes | . | . | . | Yes | . | . | Yes | . | -97.5% | 0.06 |
| c.889G>A | p.Gly297Arg | 4 | Yes | Yes | Yes | 31mth | Yes | Yes | . | Yes | . | -96.1% | 0.06 |
| c.913G>A | p.Ala305Thr | 4 | Yes | . | . | . | Yes | . | Yes | Yes | Yes | -84.5% | 0.04 |
| c.1070C>T | p.Ala357Val | 4 | Yes | Yes | Yes | 17mths | Yes | . | . | Yes | . | -90.6% | 0.04 |
| c.1084G>A | p.Gly362Arg | 4 | Yes | . | Yes | 48mths | Yes | . | . | Yes | . | -81.1% | 0.06 |
| c.1648G>A | p.Gly550Arg | 4 | Yes | . | . | . | Yes | Yes | . | Yes | . | -97.4% | 0.06 |

Taken altogether, our data suggest that variants clustered within the binding pocket of GAT-1 can properly traffic to the membrane but have poor GABA uptake due to a disruption in substrate binding mechanics (Group 3). Additionally, those variants that are buried within the inner surfaces of the protein are more likely to impact protein trafficking to the membrane (Group 2) over variants found on the outer surface of the protein (Group 1), which have minimal impacts on GABA uptake or protein trafficking. As an additional metric, protein stability algorithms deepDDG and DynaMut were performed on GAT-1 and their score outputs were analyzed against our functional data; we did not observe such correlations (**Table S2**).

### Prenatal lethality, gene-level mutability, and dominant negative effects do not contribute to missense enrichment

Our functional data strongly suggest that loss-of-function effects underlie seizures, developmental delay, and ASD phenotypes, excluding the possibility of undiscovered gain-of-function variants and complex genotype-phenotype relationships as explanations for the enrichment for missense variants observed in *SLC6A1*. We next considered whether PTVs might increase the rate of prenatal lethality leaving a disproportionate number of missense variants in cohorts of children with neurodevelopmental delay. Adjusting for gene length and DNA sequence, we find the rate of *de novo* PTVs in *SLC6A1* is equivalent to that of other NDD-associated genes (**Fig. S14A**), suggesting it is an excess of *de novo* missense variants rather than a deficit of *de novo* PTVs driving the enrichment. However, the predicted mutability of *SLC6A1* for both missense and PTV variants is equivalent to that of other NDD genes (**Fig. S14B**). As further evidence against prenatal lethality, the diagnostic rates of seizures, developmental delay, and ASD are similar for *de novo* PTVs (92%, 60%, 100% respectively) and *de novo* missense (81%, 53%, 98% respectively), furthermore, other genes associated with neurodevelopmental delay have a greater impact on early developmental milestones^39^.

We next considered whether missense variants might lead to an ascertainment bias driven by more severe symptoms due to a dominant negative effect, as observed in *SLC30A2*^16^. We tested this hypothesis by assessing co-transfections of wildtype and missense variants for two missense variants with typical GABA uptake (p.Asp43Glu, p.Ile434Met) and five severe loss-of-function missense variants (p.Gly63Ser, p.Tyr140Cys, p.Ser295Leu, p.Leu547Arg, p.Gly550Arg). None of the variants showed evidence in keeping with dominant negative effects (**Fig. S15**).

### GAT-1 vulnerability to missense variations explains missense enrichment

The GAT-1 transporter is a highly specific, dynamic, and complicated molecular machine^17^. Could the intricate protein structure render the protein unusually susceptible to loss-of-function missense variation? Exploring this possibility would require extrapolating the GABA uptake results from the 180 assayed missense variants to all possible missense variants for the gene. Multiple algorithms have been developed to estimate the functional impact of missense variants, largely based on conservation across species and constraint in human populations. We used annoVar to annotate the 180 missense variants against the output of multiple such algorithms (**Table S2**) and stepwise linear regression (**Table S6**) and random forest machine learning (**Table S7**) to build a predictive model. The R^2^ was above 0.55 for three linear regression models independently, ClinPred: R^2^=0.58, P=7×10^-35^ (**Fig. 4A**)^40^, MetaSVM: R^2^=0.57, P=8×10^-34^ [^41^], and MetaRNN: R^2^=0.55, P=1×10^-32^ [^42^]), of note, all three are ensemble models integrating multiple other algorithms trained against large databases, such as ClinVar. Selecting ClinPred and adding additional predictors (MetaSVM^41^, fitCons^43^, phyloP30way mammalian^44^, LIST-S2^45^) only marginally improved the prediction to R^2^=0.62, probably because of the high correlations between these algorithms (**Fig. S16**). Using a random forest approach to model fitting for the 180 missense variants yielded an R^2^=0.55. Given the risk of overfitting to a relatively small dataset, we elected to use the ClinPred model with linear regression alone (scaled from no predicted impact at 0 to severe predicted impact at 1).

**Figure 4.**
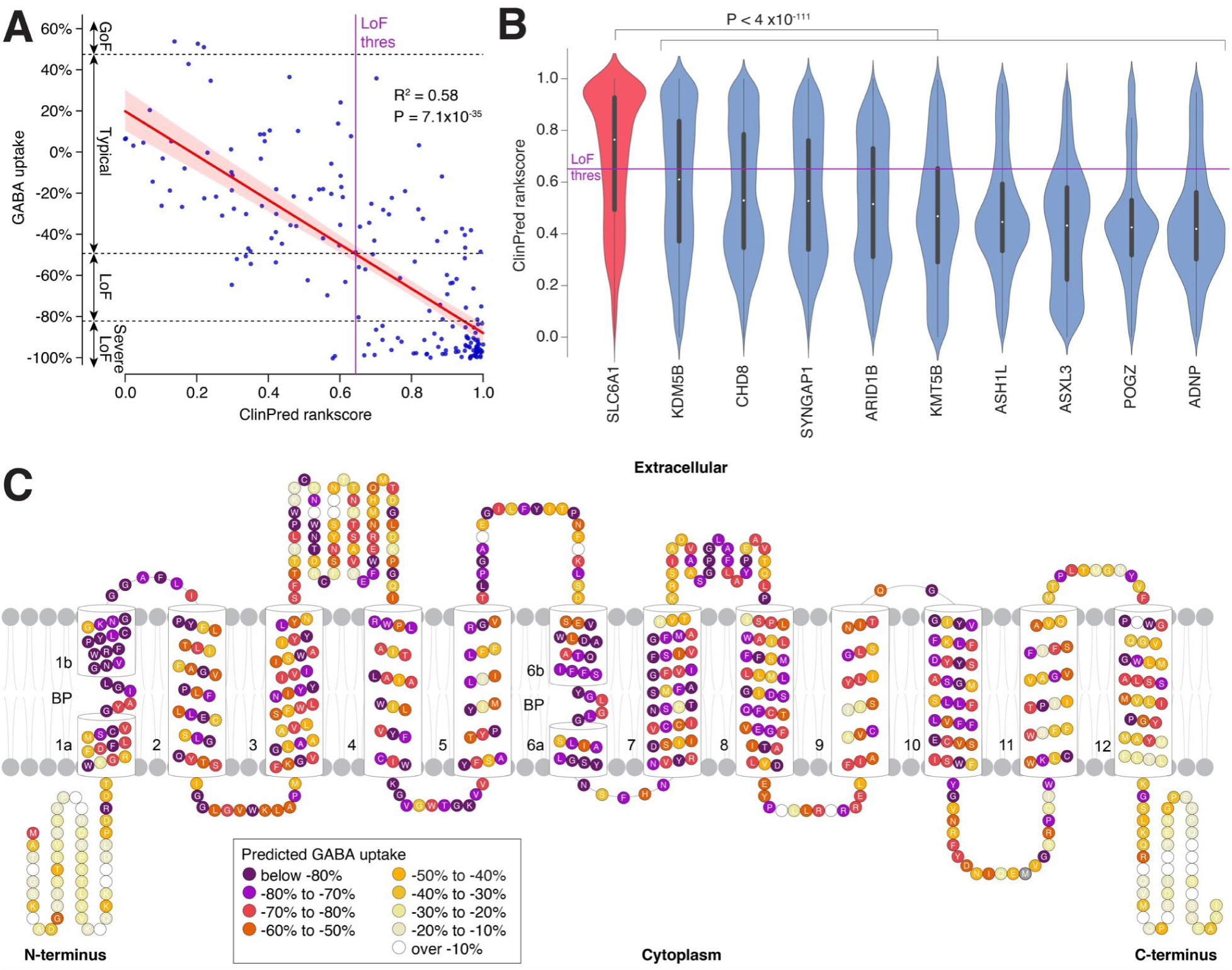
GAT-1 vulnerability underlies the enrichment of SLC6A1 missense variants. **A)** Correlation of individual GABA uptake data and ClinPred rankscore for 180 *SCL6A1*/GAT-1 missense variants. The red line shows the linear regression model with 95% confidence intervals represented by the red shading. The ClinPred value at the estimated loss-of-function threshold is indicated by the purple line (‘LoF thres’). **B)** Violin plots represent the ClinPred rankscore distribution for all possible missense variants in SLC6A1 genes and nine equivalent PTV-enriched ASD- and NDD-associated genes. The ‘LoF thres’ line from ‘A’ is also shown. **C)** All 599 amino acids of GAT-1 are colored by the mean predicted GABA uptake of all possible missense variants using the linear regression model from ‘A’ from the annotated ClinPred rankscores. Abbreviations: GoF: gain-of-function, LoF: loss-of-function. Statistical tests: A: linear regression, B: two-sided Wilcoxon test.

To select comparator genes, we identified nine genes with equivalent evidence of association with ASD (FDR ≤ 1×10^-10^) and NDD (FDR ≤ 1×10^-10^) that were enriched for PTVs (*de novo* PTV count / *de novo* missense count > 2)^2^ and had ClinPred scores available. For *SLC6A1* and these nine genes, we used the longest protein-coding isoform from GENCODE v33 to predict all possible missense variants and plotted the ClinPred score distributions (**Fig. 4B**). In keeping with a higher vulnerability to missense variation, ClinPred scores are substantially higher for *SLC6A1* than for the other nine genes, with a median of 0.76 for *SCL6A1* vs. 0.60 for *KDM5B* (P=4×10^-111^), the next highest gene (**Fig. 4B**). In NDD, we observed a 7-fold enrichment of *de novo* missense variants compared to *de novo* PTVs (43 missense, 6 PTVs, ratio=7.17)^2^. Based on a ClinPred score of ≥0.642, set by the loss-of-function threshold (**Fig. 4A**), we observed the total number of predicted loss-of-function missense sites to be 7-fold higher than the total number of predicted PTV missense sites (2,481 missense, 349 PTVs, ratio=7.11, **Table S7**), with a similar ratio after adjusting for mutability (ratio=7.06), suggesting the magnitude of this missense vulnerability is sufficient explain the observed missense enrichment. Other measures of missense severity highly correlated to the functional data yield similar patterns (**Fig. S17**).

Using the linear model to convert the ClinPred scores to estimated GABA uptake, we plotted the mean functional impact for each of the 599 amino acids in GAT-1 (**Fig. 4C**). In keeping with the 3D structure results, we see substantial vulnerability throughout transmembrane domains 1 to 10, along with some of the cytoplasmic and extracellular loops. In contrast, the N- and C-terminus loops show limited vulnerability to missense variants, consistent with our directly assayed functional data (**Fig. 1A**).

### Recurrent missense variants in *SLC6A1* are at hypermutable sites

GAT-1 vulnerability can account for the enrichment of missense variants in *SLC6A1*, however, this still does not account for the observation of recurrent *de novo* missense mutations (**Table 1**, **Fig. 2, S1**). To assess whether mutability may be a factor, we estimated the mutability of all possible missense variants in *SLC6A1* and plotted these estimates against the predicted GABA uptake from ClinPred (**Fig. 5**). A small fraction of missense variants in *SLC6A1* (75 out of 3,943, 1.9%) are at CpG sites that are known to be hypermutable^46^ due to the spontaneous deamination of 5-methylcytosine (5mC) to thymine (**Fig. 5**). However, 22 of the 31 recurrent missense variants (71.6%) and all 14 (100%) of the missense variants identified in three or more individuals are in this small hypermutable subset (**Fig. 5, S18**). The majority of these are at sites we predict to be loss-of-function.

**Figure 5.**
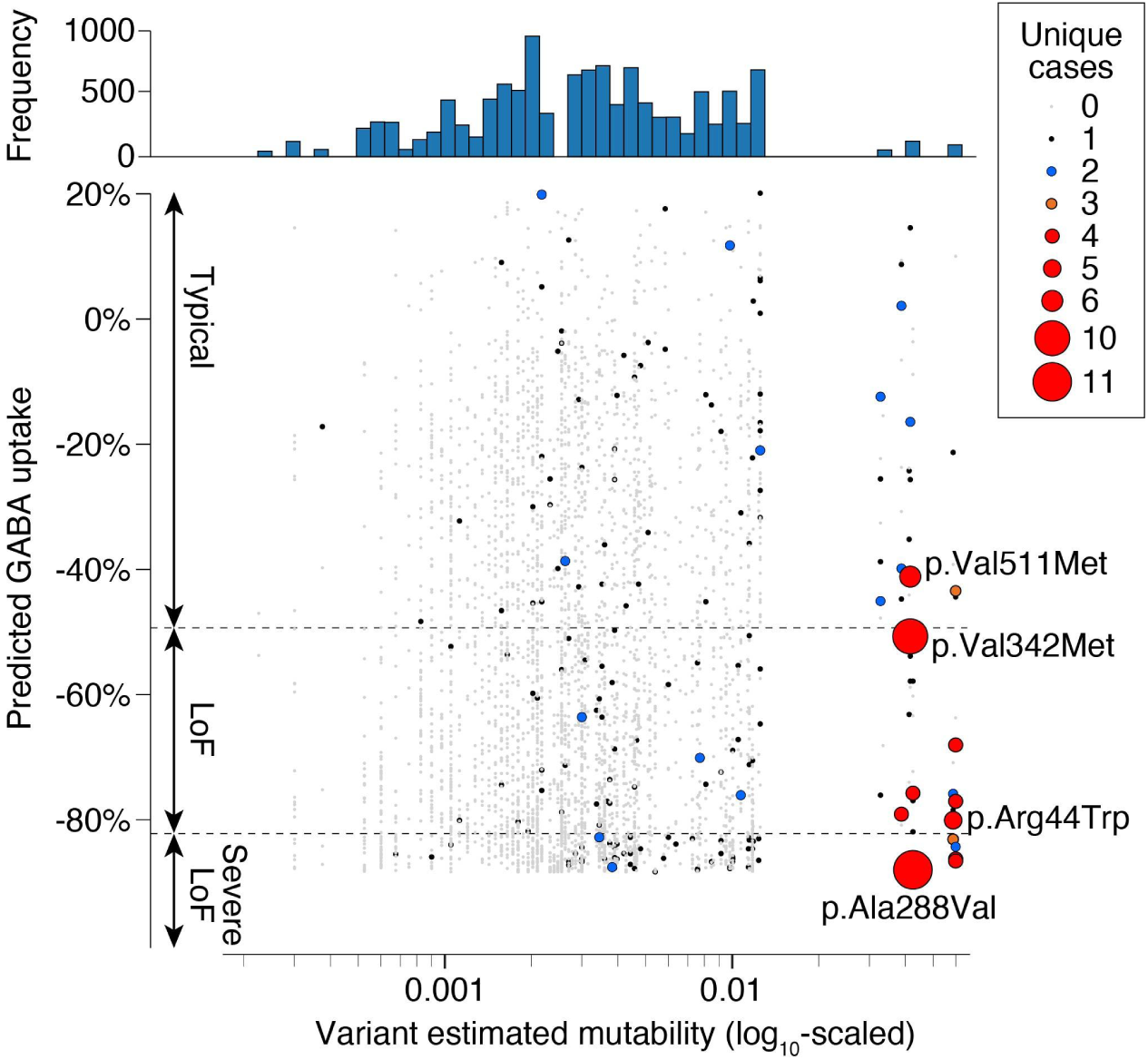
Recurrent missense variants in *SLC6A1* are at hypermutable loci. Estimated mutability based on three base-pair DNA sequences is shown for all 3,952 possible missense variants in *SLC6A1* (x-axis). A small number of variants have a ∼10-fold higher mutation rate due to the presence of CpG, as shown by the histogram (top). Predicted GABA uptake is predicted for each of these missense variants based on ClinPred (Fig. 4). The number of unique cases is indicated by size and color for each variant; the four most frequent are labeled (Fig. 2 and Table 1). Abbreviations: LoF: Loss-of-function.

## Discussion

*SLC6A1* is a promising therapeutic target due to its strong association with neurodevelopmental disorders and seizures and the clear role of the encoded GAT-1 protein in transporting GABA. However, translational progress requires a clear understanding of genotype-phenotype relationships. Some patients have protein-truncating variants (PTVs), suggesting autosomal dominant loss-of-function (haploinsufficiency, i.e., insufficient GABA transport) as the mechanism, however, the vast majority of variants have a missense effect. This enrichment of missense variants, combined with multiple recurrent missense mutations, implies a gain-of-function or dominant-negative mechanism (i.e., some missense variants have a greater impact on phenotype than PTVs). The application of a therapy designed to potentiate GABA uptake in a patient with a gain-of-function variant could have devastating consequences.

By performing the largest functional screen for *SLC6A1* to date, we see clear evidence that a reduction in GABA uptake underlies neurodevelopmental and seizure symptoms. Of the 66 germline *de novo* variants assessed, 64 (97.0%) resulted in loss-of-function (**Fig. 2E**) and the remaining two showed a trend towards reduced GABA uptake (−20.2% for p.L366V, −41.7% for p.W495L). This consistent functional result, alongside the observation that seizures, developmental delay, and autism spectrum disorder are often co-morbid in the same individual, supports a common mechanism of reduced GABA uptake leading to seizures. Associated seizure types include EMAS, EOAE, CAE, DEE, and LGS but not NAFE (**Fig. S8**). Of note, no association was seen between the age of seizure onset and the degree of GABA uptake impairment (**Fig. S8**) and other metrics of phenotypic severity were not available for assessment. While the results are suggestive that reduced GABA uptake contributes to schizophrenia risk (**Fig. 2E, S8**), further genomic, functional, and longitudinal phenotyping will be required for definitive answers.

In contrast to loss-of-function, we find limited evidence for phenotypic effects from variants that increase GABA uptake (e.g., gain-of-function). Nine missense variants were estimated to increase in GABA uptake by ≥20% (**Fig. 1**), however, analysis of ten variants using stable transfections, a more accurate method, showed more modest changes, ranging from 8% to 28% over wildtype (**Table S4**). While several of these variants that modestly increase GABA uptake are identified in patients with seizures, none of the patients are reported to have EMAS, none are classified as pathogenic/likely pathogenic in ClinVar, none are known to be *de novo*, and most are observed in the general population (gnomAD). Based on these data, the neurodevelopmental risk mediated by these variants is probably very small, if any. We considered the possibility that increased GABA uptake might have a protective effect. Genome-wide association studies (GWAS) for epilepsy do not identify significant association at the *SLC6A1* locus^47^, however, the allele frequency of these variants remains below the GWAS detection threshold of ∼2% AF.

While the functional data provide a clear answer for the impact on GABA uptake, they do not account for the observed enrichment of missense variants compared with other neurodevelopmental genes; failure to explain this phenomenon could reduce confidence in future therapies aiming to increase GABA uptake. Co-transfection of wildtype and missense variants did not find evidence of dominant negative effects (**Fig. S15**) and genomic and phenotype data do not support prenatal lethality from PTVs. In contrast, using computational predictors of missense severity to extrapolate the functional data to all possible missense variants provides a clear answer: *SLC6A1*/GAT-1 is simply more sensitive to disruption by missense variants than many genes. This observation also explains the relatively high frequency of *SLC6A1*-related disorders in large cohorts despite the comparatively small size of the gene (599 amino acids). While missense sensitivity can explain the missense enrichment, at face value it does not account for the recurrent missense variants. However, integrating mutability data, we observe that these can be explained by a small number of missense variants that both reduce GABA uptake and occur at highly mutable CpG sites. This missense sensitivity may also explain other genes enriched for missense variants despite predominantly loss-of-function effects, such as *SCN1A* (Dravet Syndrome).

Our functional data provide ‘Strong’ information to guide clinical interpretation based on the ACMG Guidelines (specifically, categories PS3 and BS3). Of the 213 variants we tested, 153 (71.8%) did not have ACMG classifications (e.g., Likely Pathogenic) in the ClinVar database (**Fig. 2D**). Considering the 127 we identified as loss-of-function or severe loss-of-function, only 30 (23.6%) had previously been classified as Pathogenic or Likely Pathogenic with the remainder being unscored (66, 52.0%), Uncertain Significance (18, 14.2%), Conflicting Interpretations (10, 7.9%), or Likely benign (3, 2.4%). In contrast, of the 86 variants that did not show significant loss-of-function, only 15 (17.4%) were classified as Benign or Likely Benign, with the remainder being unscored (35, 40.7%), Uncertain Significance (28, 32.6%), Conflicting Interpretations (6, 7.0%), Likely Pathogenic (1, 1.2%), or Pathogenic (1, 1.2%,) (**Fig. 2D, Table S2**).

Overall, for the 60 variants that were scored, only three had opposing clinical and functional interpretations, recorded as Likely Benign in ClinVar but observed to be loss-of-function for GABA uptake: p.Val142Ile (−50.4% GABA uptake), p.Arg417Cys (−63.5%) and p.Val464Glu (−61.5%). All three variants were submitted to ClinVar by the same submitter without additional evidence for the variant being benign. The associated condition was EMAS/MAE and therefore consistent with *SLC6A1* haploinsufficiency. The clinical interpretation was not supported by the functional data for two further variants: p.Leu366Val (Likely Pathogenic, −20.2% GABA uptake, not in gnomAD, germline *de novo* in an individual with developmental delay but no further clinical details) and p.Val323Ile (Pathogenic, −30.0%, rare in gnomad (7×10^-6^), unknown inheritance in an individual with developmental delay but no further clinical details). We note that our loss-of-function threshold of −49.2% is probably conservative and that milder impairments may be clinically relevant, especially on a sensitized genetic background.

To gain further insight into the functional impact of *SLC6A1* variants causing these phenotypes, we studied whether there were any structure-functional relationships. Previous studies have shown that mutations in SLC6 family members, including *SLC6A1*, can cause a disruption in protein trafficking to the membrane, and thus impact function and lead to disease^10,12,48,49^. Our study demonstrated a broad clustering of functionally studied variants by whether or not they were detected on the plasma membrane through high-content imaging of GFP expression. We showed that variants clustered in three distinct groups (**Fig. 3A**): Group 1) variants present on the cell surface with typical uptake (n=30), Group 2) variants absent from the cell surface with low uptake (n=40), and Group 3) variants that were present on the cell surface but had low uptake (n=16). Notably, Group 3 variants that showed proper protein trafficking and low transporter function were clustered within the binding pocket of the GAT-1 protein and/or the interior of the 3D structure. This suggests that variants in these structural locations are disrupting proper substrate binding and/or transport mechanics (**Fig. 3D and E**). Those in Group 2 (low uptake and low cell surface expression) shared similar mapping on the 3D structure to Group 3, found mainly in positions embedded within the inner surface of the protein and on transmembrane domains (**Fig. 3D, Fig. S12B, and S12D**). This shows the importance of those regions, most notably the transmembrane domains, in proper transporter function and perhaps stability and proper folding before trafficking.

Understanding where mutations are structurally located and whether they impact trafficking is important for the design of future therapies. For example, before proteins can traffic to their destination, they must properly fold post-translation and in mutants, this folding can be disrupted. In this case, a therapy that increased the expression of both alleles might have unexpected consequences. Certain SLC6 family members have been previously rescued from folding-deficient and disease-associated mutations through the use of pharmaco-chaperoning^50^. To date, GAT-1 has not been studied enough to understand whether using chaperones could help improve symptoms. Additionally, while we broadly studied the structure-functional relationships of a subset of variants, many other pathways influence transport and trafficking mechanisms of GAT-1. This includes N-glycosylation, oligomerization disruption, protein kinase C-mediated phosphorylation, endocytic trafficking, and more^49,51–55^. With the latest advances in GAT-1 structure, new studies need to be conducted to better understand how GAT-1 works in cells mechanistically.

As ever, our experiment has limitations. The functional studies were performed in HEK cells rather than the cell types in the brain that mediate symptoms. Also, we only assayed GABA uptake; gain-of-function consequences could include the uptake of another molecule or an entirely orthogonal function altogether. However, gain-of-function seems unlikely, given the specificity of the transporter to GABA, the functional enrichment of transmembrane domains, the limited evidence of phenotypic consequences of variants in the N- and C-termini, and the observation that missense sensitivity and mutability can account for the missense enrichment.

## Conclusion

In summary, by integrating genomic, functional, and phenotype data, we observe unambiguous evidence that reduced GABA uptake underlies neurodevelopmental sequelae associated with *SLC6A1* variants, including seizures, developmental delay, and autism spectrum disorder. Furthermore, the observed enrichment of missense variants and recurrent missense variants can be explained by the sensitivity of *SLC6A1*/GAT-1 to missense variants and mutable CpG sites respectively. Based on these results, therapeutic strategies for *SLC6A1*-related neurodevelopmental disorders should aim to increase GABA uptake.

## Supporting information

Supplemental Figure S13 (MPG)

## Data Availability

All data produced in the present work are contained in the manuscript and the associated supplementary files.

## Acknowledgements

The BioMarin group wants to thank Amber Freed of SLC6A1Connect, Prof. Dennis Lal, and Prof. Jing-Qiong Kang for reviewing the experimental design. This study incorporates data generated by the DECIPHER community. A full list of centers who contributed to the generation of the data is available from https://deciphergenomics.org/about/stats and via email from. Funding for the DECIPHER project was provided by the Wellcome Trust. We used the Invitae Explorer to explore variants and aggregate testing data from Invitae patients, including genomic and demographic data. We thank Alliance Pharma for their support developing and running the BLA and GABA mass spectrometry assays.

The UCSF group would also like to extend gratitude to the SLC6A1 Connect organization and Amber Freed, and the American Epilepsy Society (Seed Grant to SJS) and the National Institute of Mental Health (R01MH116999 and R01MH129751 to SJS) for funding. We thank Michelle Arkin Lab at UCSF for the use of their IN Cell Analyzer and IN Carta software. Specifically, Jezrael La Fuente for his training, guidance and troubleshooting analysis of the high content imaging data. We are also grateful for Avner Schlessinger and Keino Hutchinson for their structural biology expertise, helping us accurately dock GABA onto the cryo-EM GAT-1 structure, and for their interpretation in regards to our data. A special thanks to Cornelius Gati for kindly sharing his GAT-1 structure annotations with us.

## Author contributions

Arthur Wuster and Steven Froelich designed and supervised the study. Marena Trinidad contributed to study design, generated constructs, and executed functional experiments. Marena Trinidad and Arthur Wuster analyzed the data. Arthur Wuster and Lorenzo Bomba, compiled the list of variants characterized by BioMarin. William Wallace performed cell based uptake assays and Geoffrey Berguig designed the mass spectrometry assays. Jon H. LeBowitz and Karol Estrada conceived the project and contributed to study design. Marena Trinidad, Arthur Wuster and Steven Froelich contributed to manuscript writing.

Stephan J. Sanders helped design and supervise the study, analyze data and wrote the manuscript. Kathy Giacomini supervised the study and revised the manuscript. Dina Buitrago Silva helped design the experimental study, performed transport uptake assays, performed high content image experiments, analyzed data and wrote the manuscript. Alicia Ljungdahl analyzed the data and wrote the manuscript. Cory Patrick assessed the random forest model and Shan Dong contributed the mutability analysis. Jezrael La Fuente supervised, provided expertise, and analyzed high content imaging experiments. Michelle Arkin provided instrumentation (IN Cell Analyzer 6500 and IN Carta Software). Avner Schlessinger provided expertise on transporter structure and interpretation of variant impact. Keino Hutchinson provided insight to variant impact on structure, DynaMut2 results and docked GABA onto the GAT-1 PDB file.

## Web resources (if any)

None

## Data and code availability

All data are reported in the supplementary tables.

