## Supplemental figures and tables for "Haploinsufficiency underlies the neurodevelopmental consequences of *SLC6A1*/GAT-1 variants"

Supplemental Table S1: Individual-level data. Excel spreadsheet in which each row represents an individual with data on the SLC6A1 variant and phenotype.

Supplemental Table S2: Variant-level data. Excel spreadsheet in which each row represents a unique variant with data on the SLC6A1 variant and phenotype combined across individuals.

Supplemental Table S3: Plasmid sequences. Excel spreadsheet with four sheets detailing the plasmids used for the GABA uptake and surface expression assays.

Supplemental Table S4: GABA Uptake and Surface Expression data. Excel spreadsheet with four sheets detailing the results of the GABA uptake and surface expression assays.

Supplemental Table S5: ClinVar Categories. Excel spreadsheet showing the relationship between ClinVar category and GABA uptake.

Supplemental Table S6: Linear Regression. Excel spreadsheet with 10 sheets detailing the stepwise linear regression model building for missense variants.

Supplemental Table S7: All SLC6A1 SNVs. Excel spreadsheet showing functional annotation, GAT-1 location, and missense severity scores (including predicted from linear regression model and random forest model) for all possible SNVs in SLC6A1 cDNA.

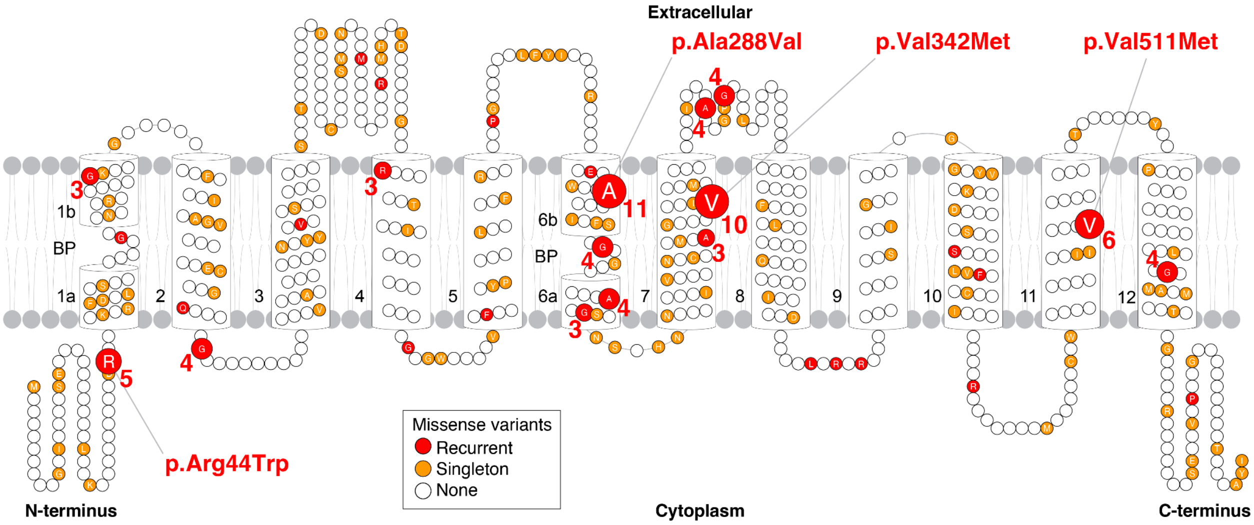


**Supplemental Figure 1. Recurrent missense variants in *SLC6A1*.** 2D representation of the 599 amino acids in the GAT-1 protein showing singleton (orange) and recurrent (red) missense variants. Missense variants identified in three or more individuals are represented by the size of the amino acid and the number next to it (Table S2).

| ClinVar category | Severe loss-of-function | Loss-of-function | Typical | Gain-of-function |
|---:|---|---|---|---|
| **Pathogenic** | 11 | 1 | 1 | 0 |
| **Pathogenic/Likely pathogenic** | 4 | 1 | 0 | 0 |
| **Likely pathogenic** | 12 | 1 | 1 | 0 |
| **Uncertain significance** | 13 | 5 | 26 | 2 |
| **Likely benign** | 0 | 3 | 3 | 0 |
| **Benign/Likely benign** | 0 | 0 | 2 | 0 |
| **Benign** | 0 | 0 | 10 | 0 |
| **Conflicting interpretations of pathogenicity** | 7 | 3 | 6 | 0 |
| **Not provided** | 1 | 0 | 0 | 0 |
| **Not listed** | 52 | 13 | 34 | 1 |

**Supplemental Table S2. Functional data by ClinVar clinical significance.**

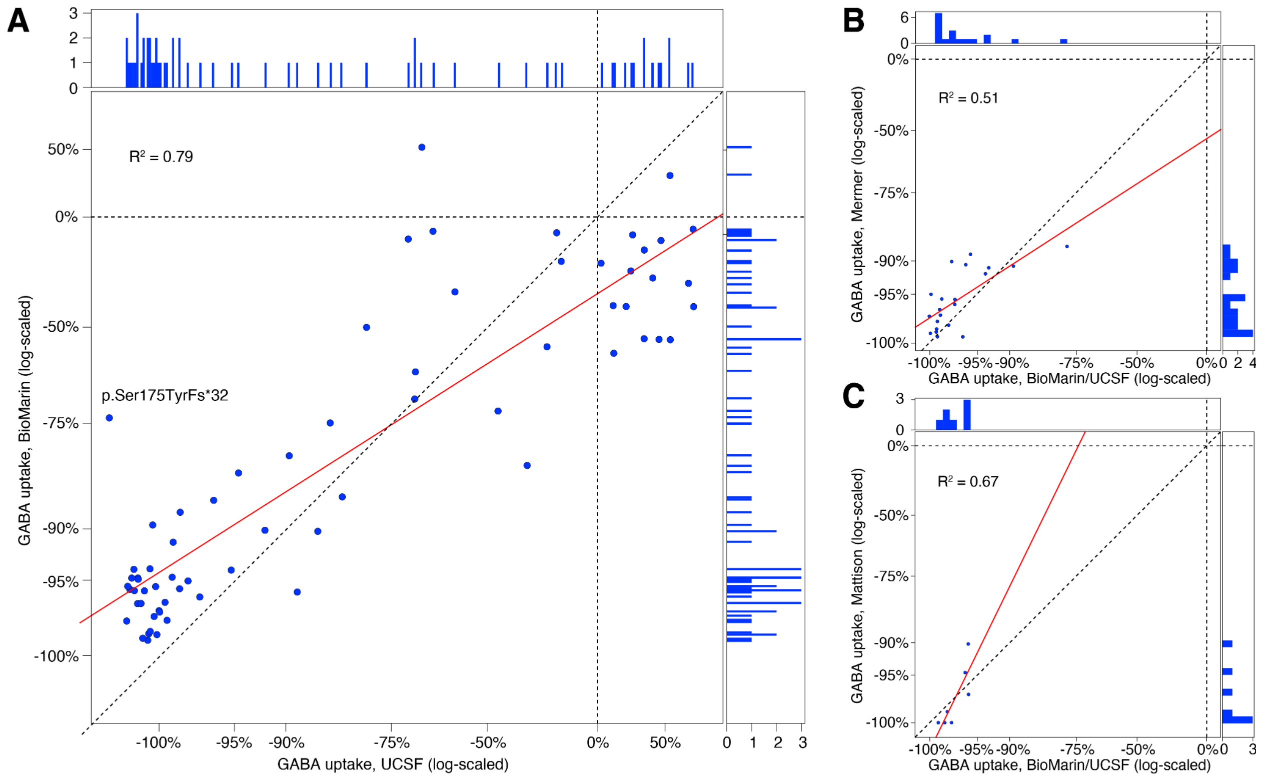


**Supplemental Figure 2. Correlation between GABA uptake data across datasets. A)** GABA uptake functional data as a percentage of wildtype generated for 68 *SLC6A1* variants assayed by both the UCSF group (x-axis) and the BioMarin group (y-axis). The red line shows the linear regression model ($R^2$ =0.79). **B)** GABA uptake data for 21 variants assayed by UCSF/BioMarin (x-axis) and Mermer group[12] (y-axis). **C)** GABA uptake data for 7 variants assayed by UCSF/BioMarin (x-axis) and Mattison group[11] (y-axis).

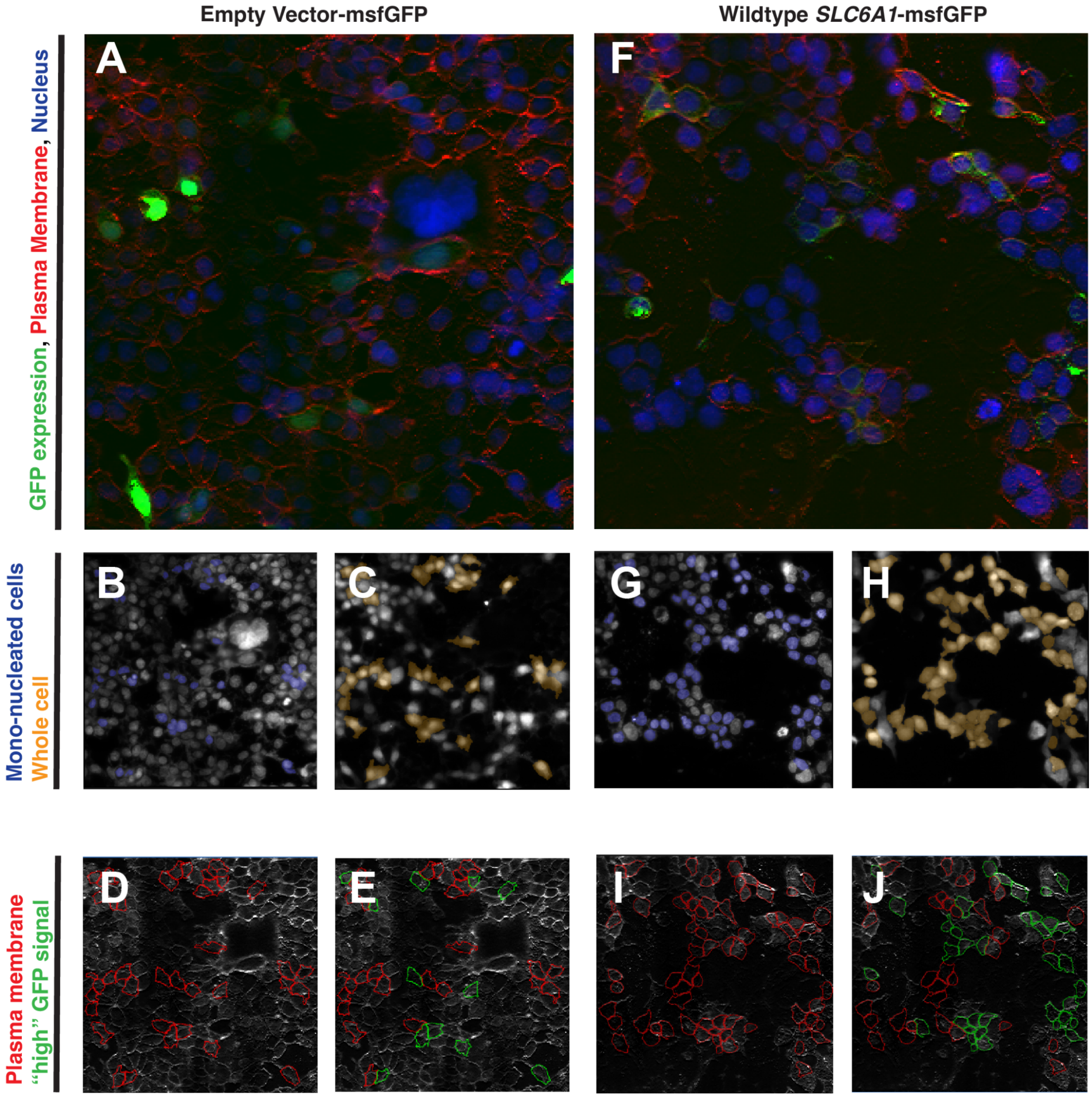


**Supplemental Figure 3: High content imaging colocalization and segmentation. A)** HEK293 cells transfected with GFP-tagged empty vector shown with colocalized fluorescence tags. IN Carta image analysis was used to identify **B)** mono-nucleated cells, **C)** whole cells, **D)** the plasma membrane, and **E)** plasma membrane in cells with "high" GFP signal. These panels are repeated for HEK293 cells transfected with GFP-tagged wildtype *SLC6A1* vector shown with colocalized fluorescence tags in **F, G, H, I,** and **J**.

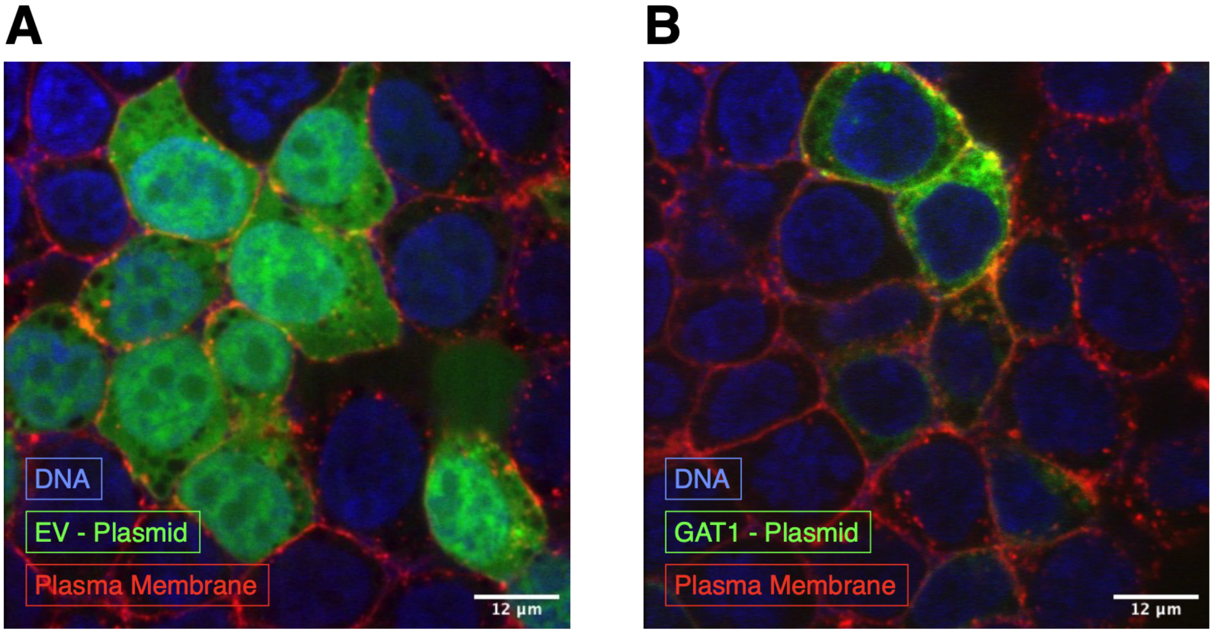


**Supplemental Figure 4. Confocal imaging of HEK293 cells.** Transient transfected HEK293 cells overexpressing **A)** empty vector GFP (EV), or **B)** GAT-1 GFP plasmid.

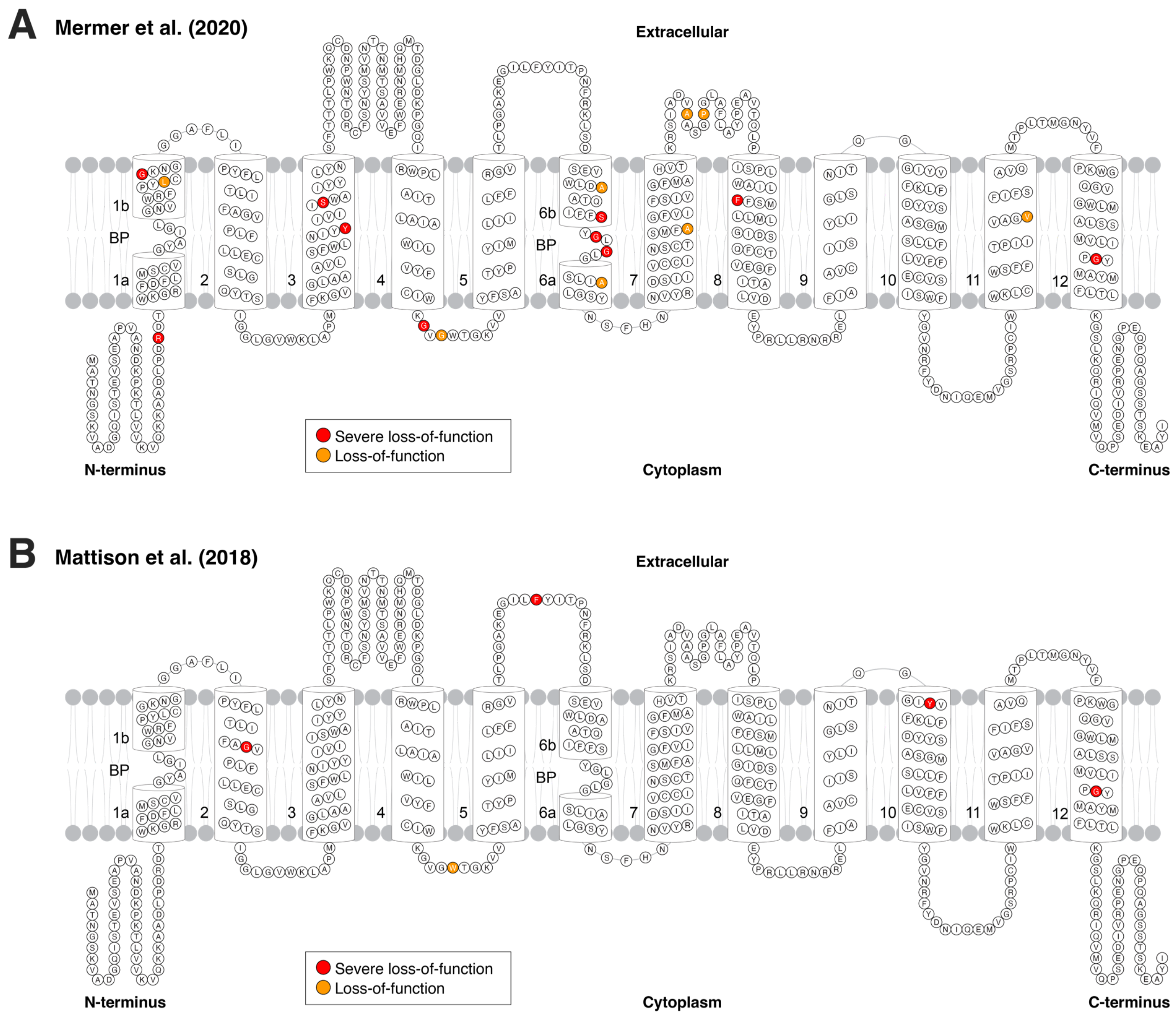


**Supplemental Figure 5. Topology of the GAT-1 protein and GABA uptake values of previous datasets.** 2D representation of the GAT-1 protein showing functional results for missense and in-frame variants from: **A)** Mermer group[12] and **B)** Mattison group[11]

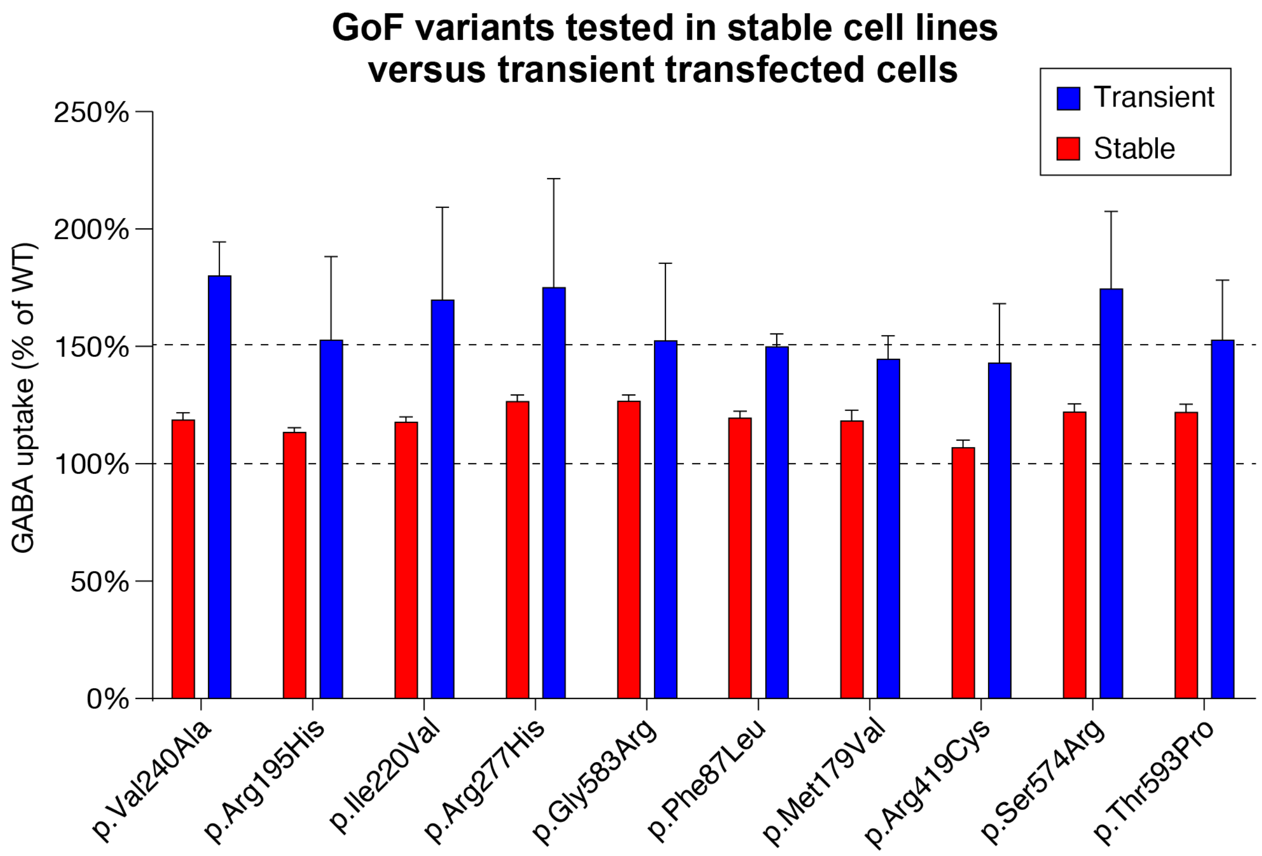


**Supplemental Figure 6: GABA uptake assay of potential gain of function variants in transient transfected cells versus stable expressing HEK293 cells.** These results (**Table S4**) are based on the highest functioning variants from UCSF data (blue bars, transient) and replicated in stable cell lines (red bars). Results are shown as a percentage of wildtype (y-axis) and error bars represent mean ± SEM of 3 biological replicates.

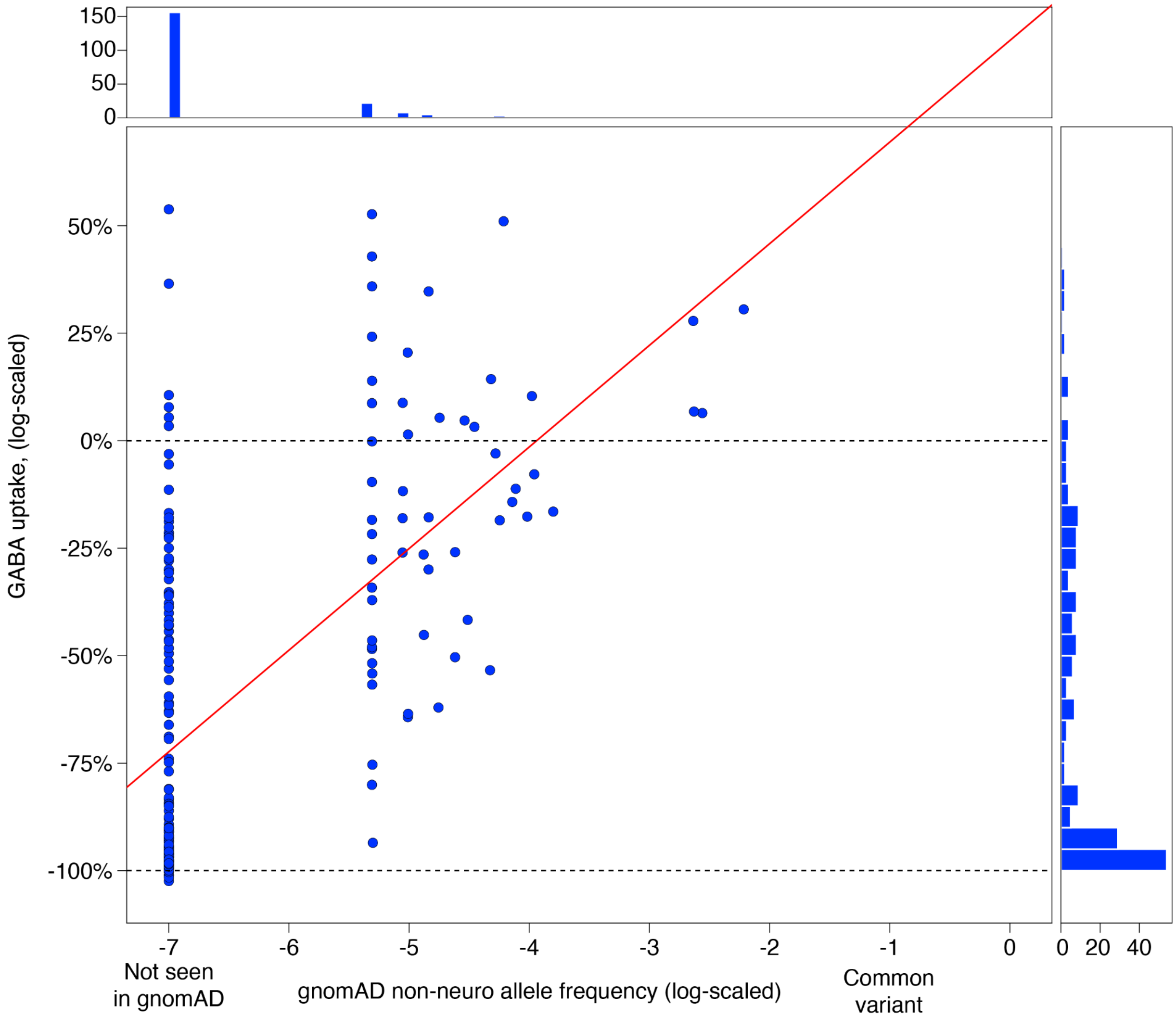


**Supplemental Figure 7. Correlation between GABA uptake and population allele frequency.** Relationship between GABA uptake functional data as a percentage of wildtype (y-axis) and population allele frequency (x-axis) based on gnomAD (v2 non-neuro) for all 213 *SLC6A1* variants assayed.

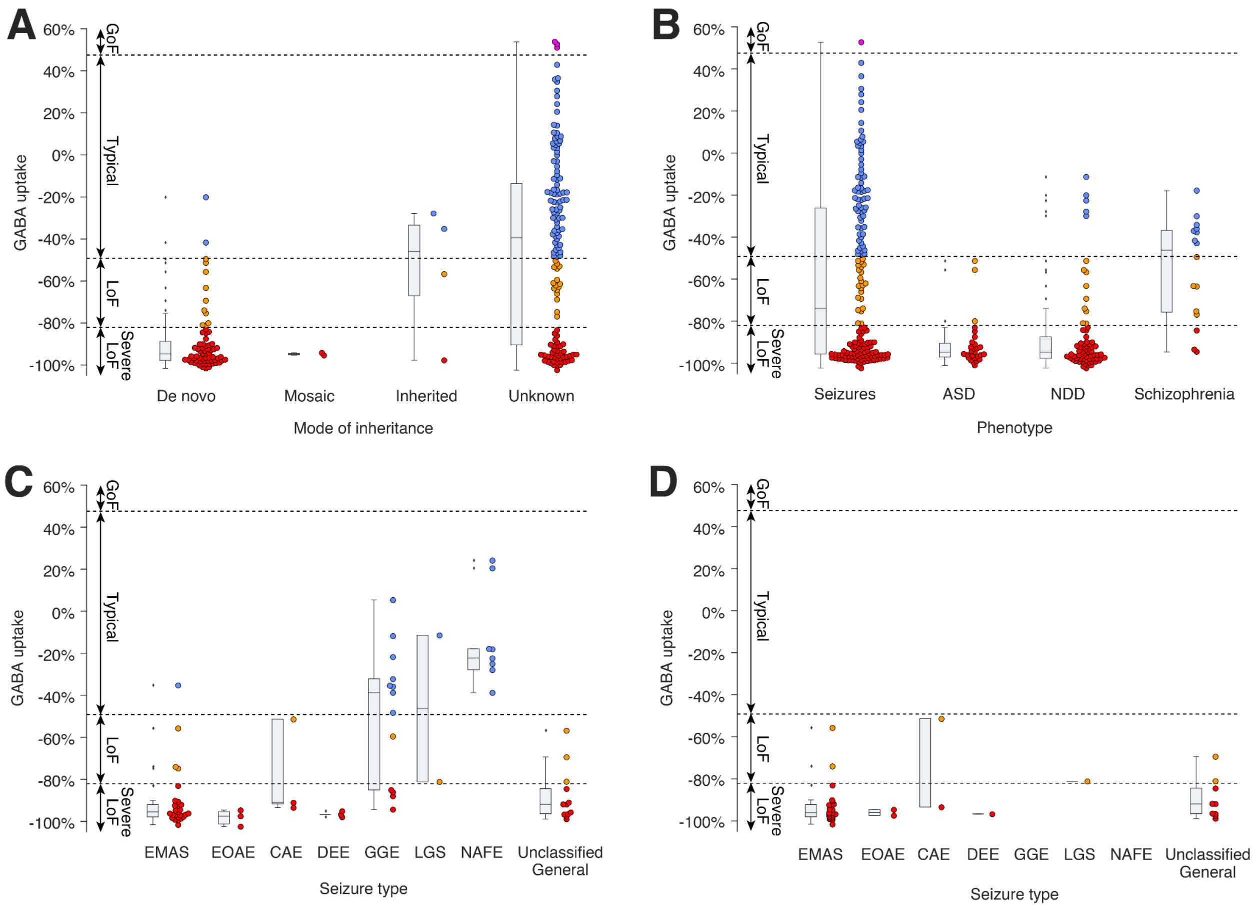


**Supplemental Figure 8. GABA uptake by inheritance, phenotype, and seizure type. A)** GABA uptake data for 213 variants by reported mode of inheritance. Each variant is only represented once; recurrent variants with multiple modes of inheritance (e.g., *de novo* and inherited) are shown in the leftmost category of any individual (**Table S1**). **B)** GABA uptake data for 197 variants by diagnosis. If multiple diagnoses are reported for the variant it is reported for all diagnoses. Sixteen variants without a diagnosis are not shown (**Table S2**). **C)** GABA uptake data for 70 variants by seizure type; each variant is assigned to only one category (leftmost if multiple seizure types). A further 93 variants were associated with seizures without a specific seizure type listed. **D)** Panel 'C' is repeated for 36 variants reported to be germline *de novo*. Abbreviations: ASD: Autism spectrum disorder, NDD: neurodevelopmental delay, EMAS: Epilepsy with myoclonic atonic seizures, EOAE: Early-onset absence epilepsy, CAE: Childhood absence epilepsy, DEE: Developmental and epileptic encephalopathy, GGE: Genetic generalized epilepsy, LGS: Lennox-Gastaut Syndrome, NAFE: Non-familial non-acquired focal epilepsy.

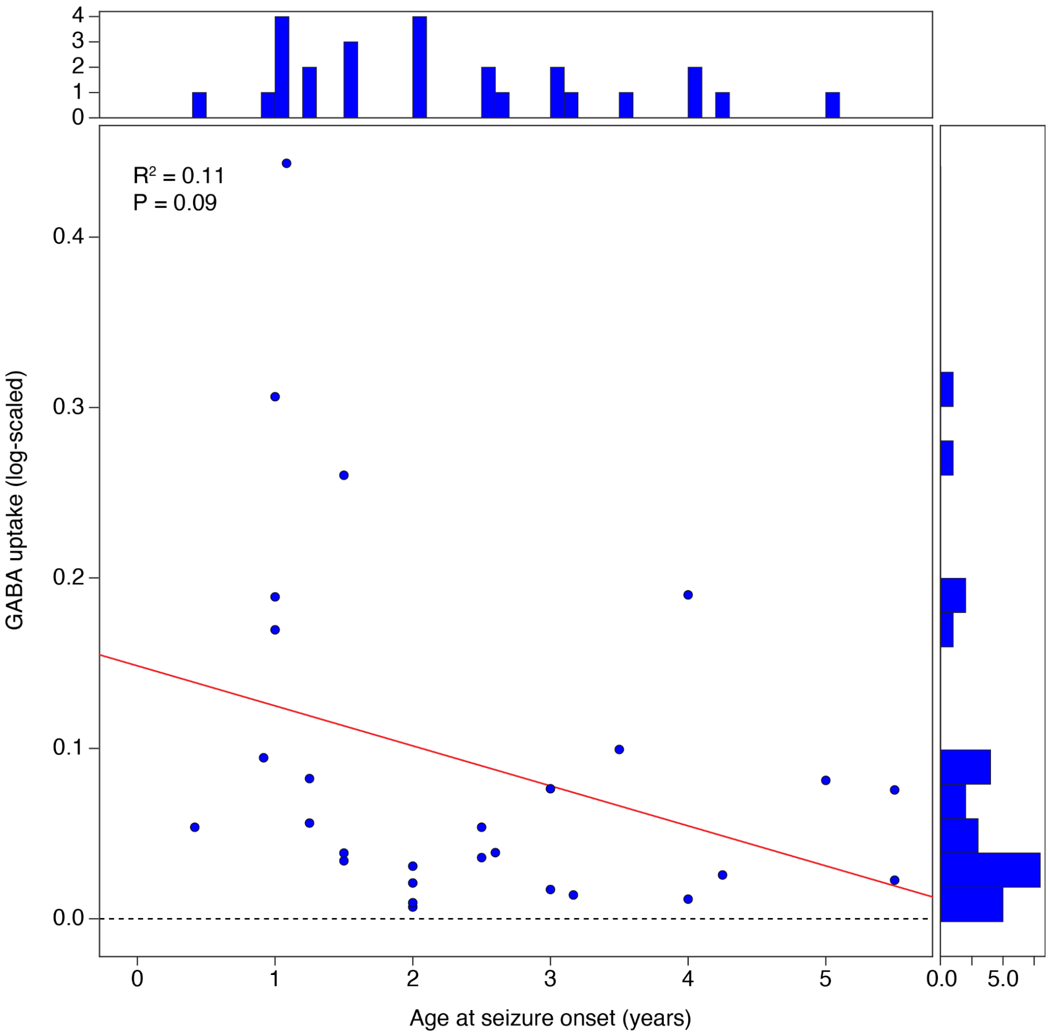


**Supplemental Figure 9. Relationship between GABA uptake and age of seizure onset.** Relationship between GABA uptake functional data as a percentage of wildtype (y-axis) and age of seizure onset (x-axis) for all 34 *SLC6A1*variants where this information was reported.

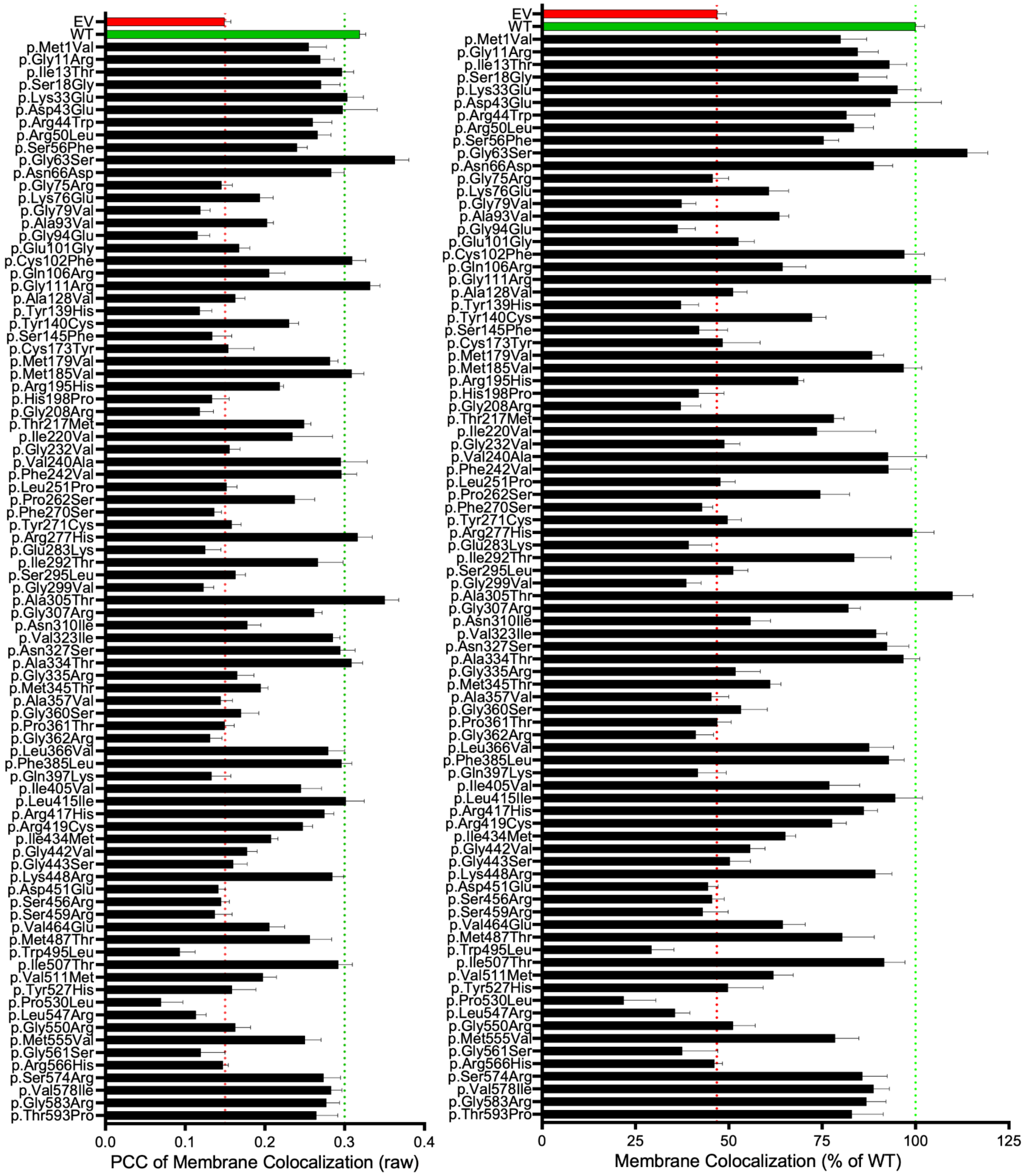


**Supplemental Figure 10: Quantitative results of high content imaging and colocalization of 86 GAT-1 variants.** Represented as **A)** Pearson Correlation colocalization (PCC) values, and **B)** as a percent of wildtype. Error bars represent mean ± SEM of 3 biological replicates performed in triplicates.

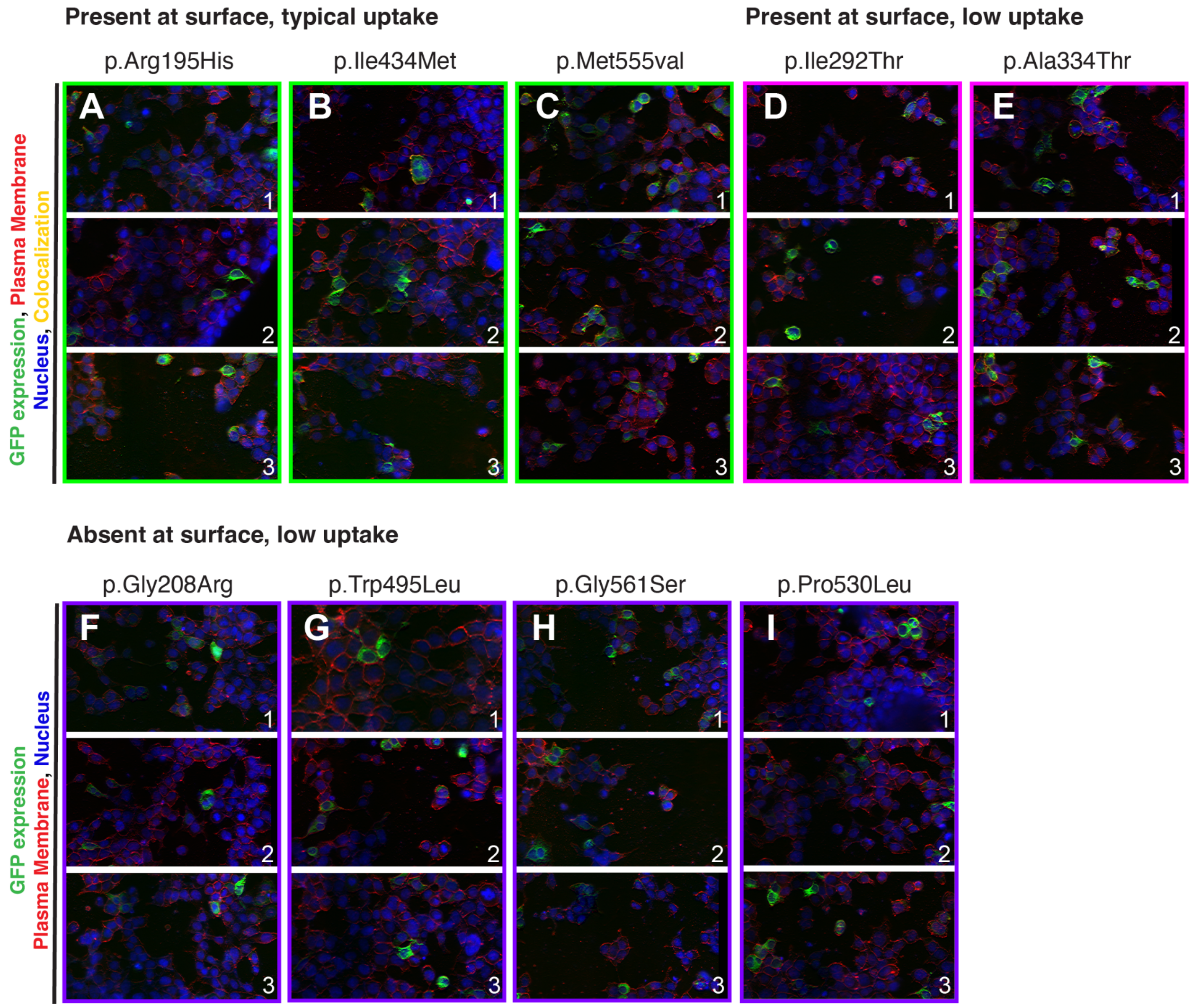


**Supplemental Figure 11: Raw images of high content imaging colocalization for a subset of missense variants.** Images are divided by clusters of GAT-1 cell surface expression presence and uptake activity and each variant is shown with three different fields of view. **A, B, C)** Present at surface and typical GABA uptake. **D, E)** Present at surface, low GABA uptake. **F, G, H, I)** Absent at surface and low GABA uptake.

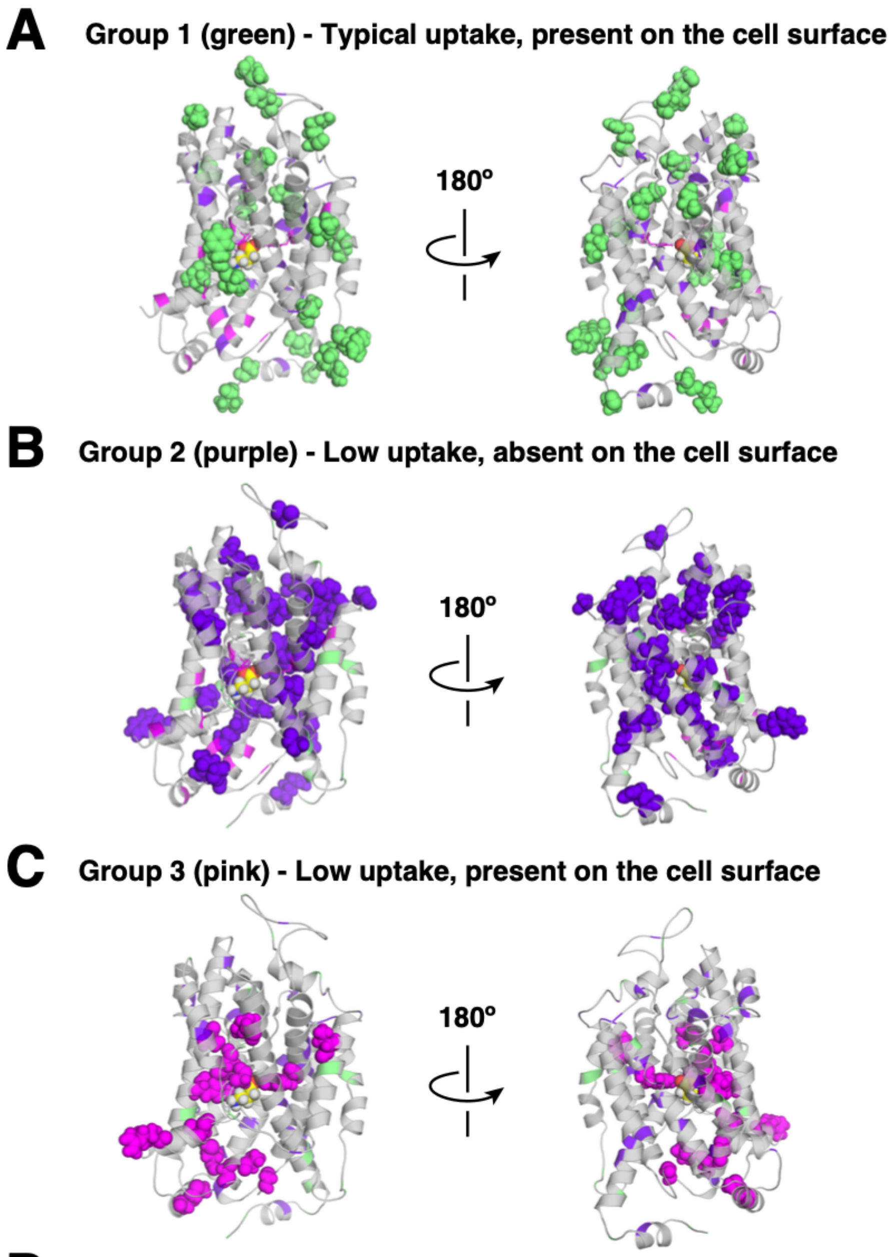


**D**

| Group | outer-surface count | inner-surface count | **outer-surface (% of total)** | **inner-surface (% of total)** |
|---|---|---|---|---|
| 1 - green | 14 | 8 | **63.6%** | **36.4%** |
| 2 - purple | 8 | 32 | **20%** | **80%** |
| 3 - pink | 1 | 15 | **6.3%** | **93.8%** |

**Supplemental Figure 12. Mapping of GAT-1 variants onto 3D structure by group of surface expression and functional type.** Variants are highlighted by group **A)** present at surface and typical GABA uptake, **B)** absent at surface, low GABA uptake, and **C)** present at surface and low GABA uptake. **D)** Individual variants mapped in A, B, and C are counted whether they appear facing at the outer-surface of the protein or the inner-surface and represented as a percent of the total within that group

**Supplemental Figure 13: PyMOL movie of GAT-1 rotating (separate file/link).**
Severe loss of function and loss of function variants are highlighted (red, orange respectively).

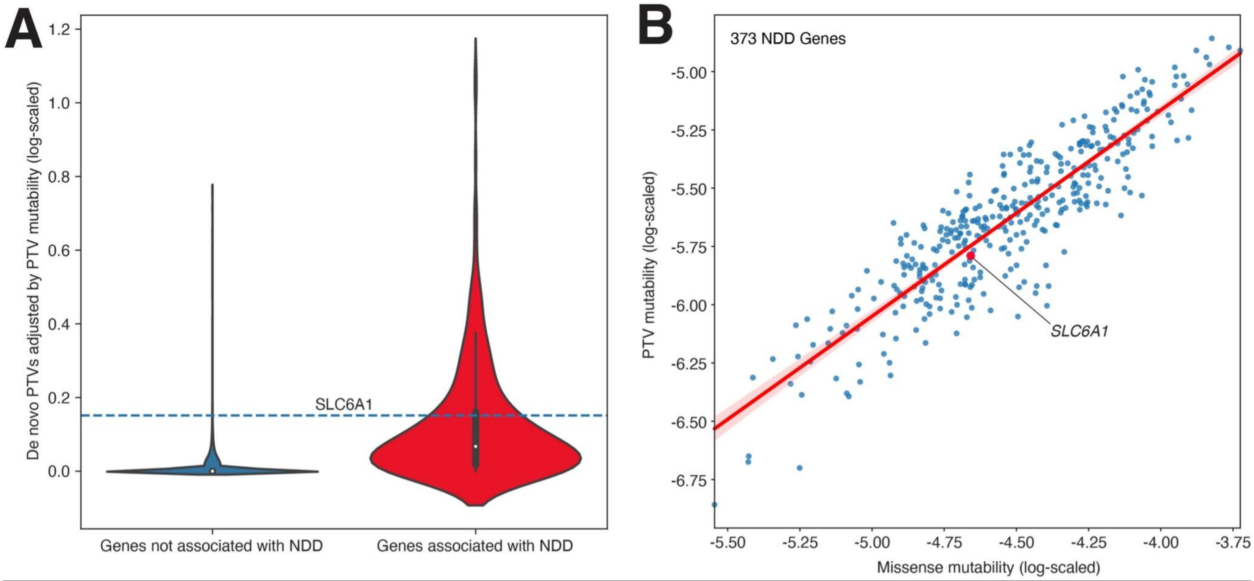


**Supplemental Figure 14: Frequency of *de novo* PTVs and gene-level mutability. A)** The observed number of *de novo* PTVs in neurodevelopmental delay (NDD) cases[2] is divided by the estimated PTV mutability rate[19]. Metrics are shown for all autosomal protein-coding genes split into 373 genes associated with NDD (red, FDR ≤ 0.001) and 16,511 genes not associated with NDD (blue, FDR > 0.1). The equivalent value for SLC6A1 is shown as the blue dashed line. **B)** The mutability is shown for 373 NDD-associated genes for missense variants (x-axis) and PTVs (y-axis).

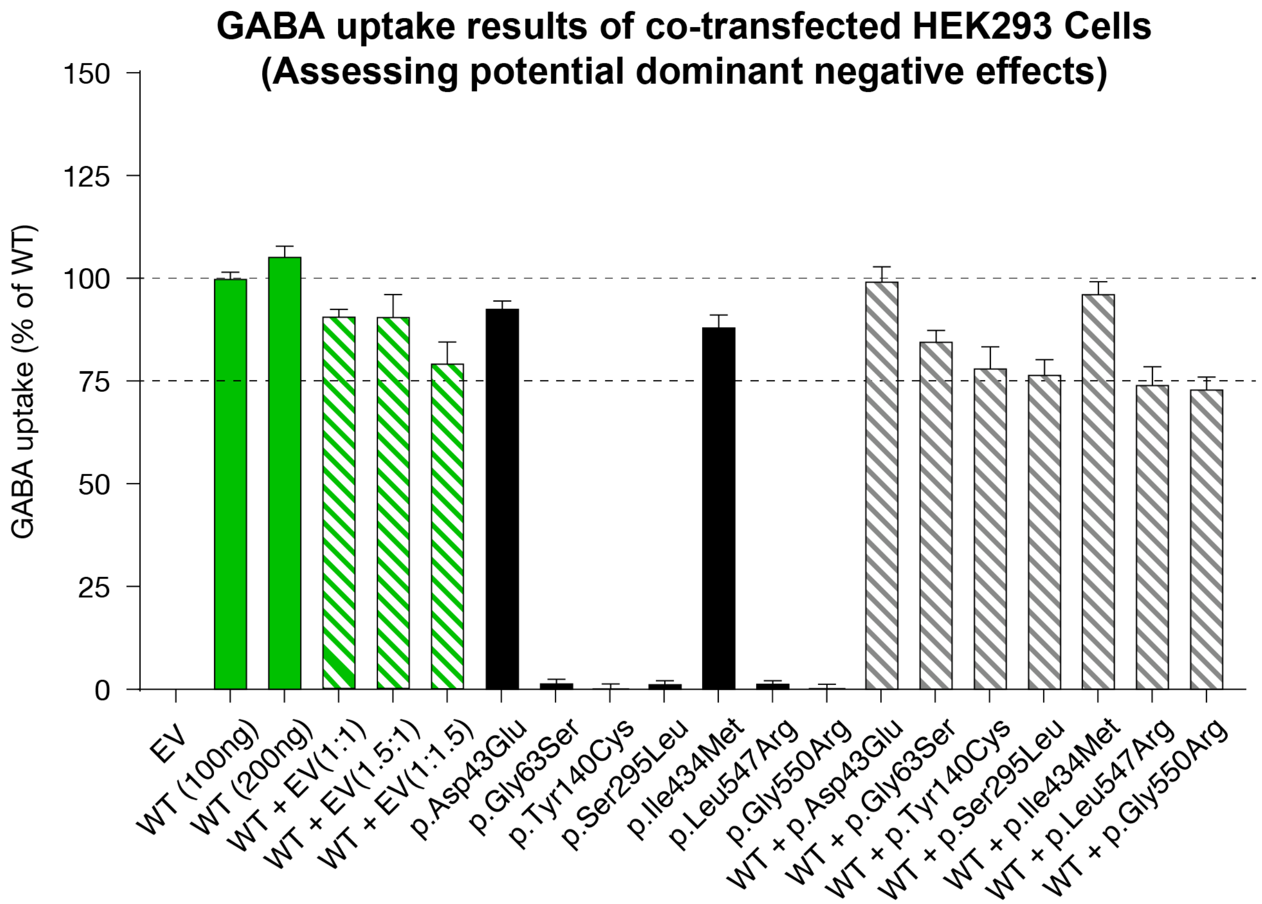


**Supplemental Figure 15: Assessing potential dominant negative effects in co-transfected HEK293 cells.** Several distinct plasmid ratios of WT to EV are shown as positive controls of co-transfection (green stripes) and WT plus variant plasmids co-transfected are at a 1:1 plasmid ratio (gray stripes). Solid green and solid black bars represent single plasmids transfected into cells at a time. Results are shown as a percentage of wildtype (y-axis) and error bars represent mean ± SEM of 3 biological replicates. No significant difference is demonstrated between co-transfected experimentals and controls (stripes).

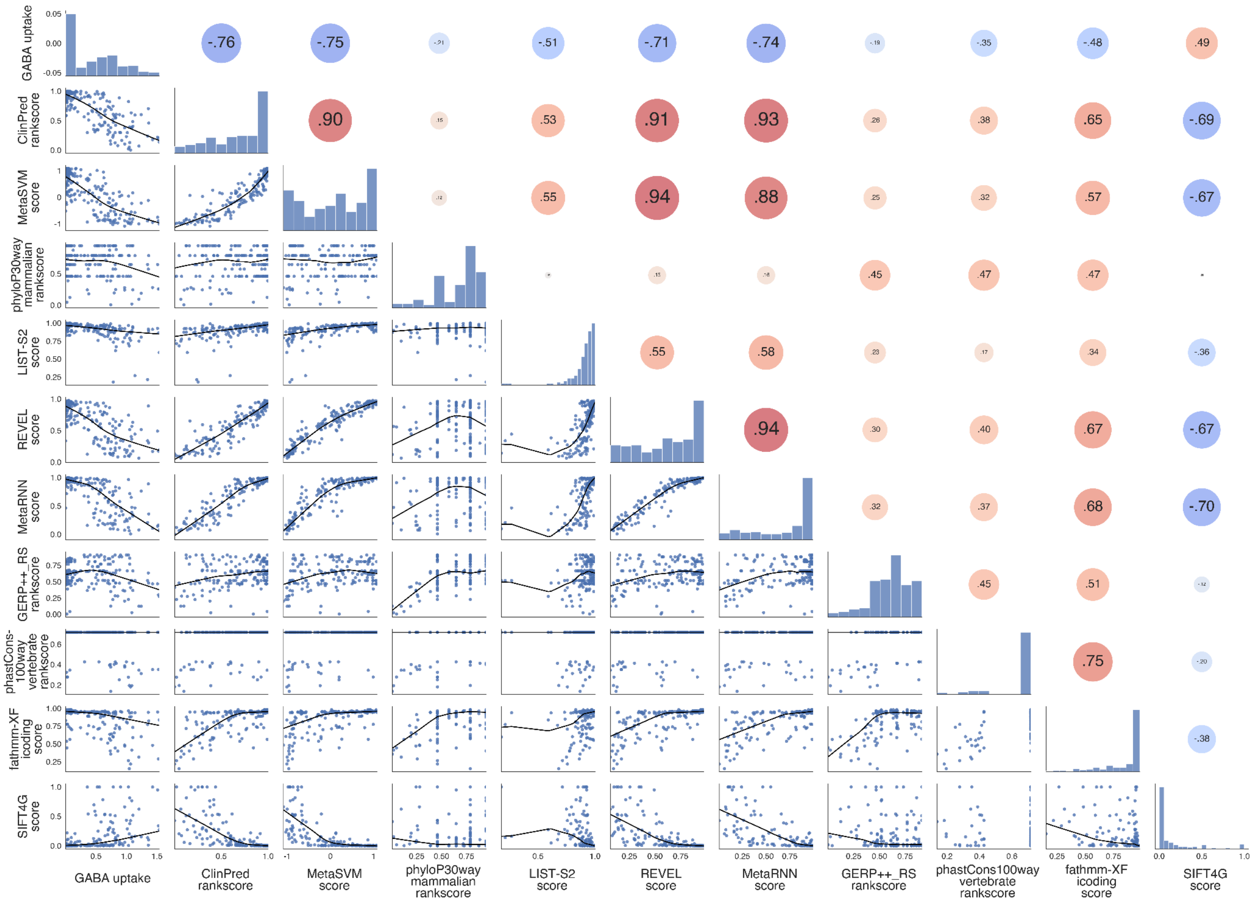


**Supplemental Figure 16: Correlation of the top ten prediction scores with GABA uptake.** Relationship between observed GABA uptake values and top ten missense severity scores from stepwise linear regression model for 178 missense variants in *SLC6A1*. Each plot represents the comparison between two out of eleven metrics (GABA uptake and ten missense scores). Plots in the bottom left show a scatter plot with a Locally Weighted Scatterplot Smoothing (LOWESS) regression curve. Plots on the diagonal (i.e., each metric against itself) show a histogram of how the metric is distributed. Plots on the upper right show the Pearson correlation (text) with the size and shade of the dot increasing with the strength of the correlation and the color representing the direction of the correlation (blue for negative, red for positive).

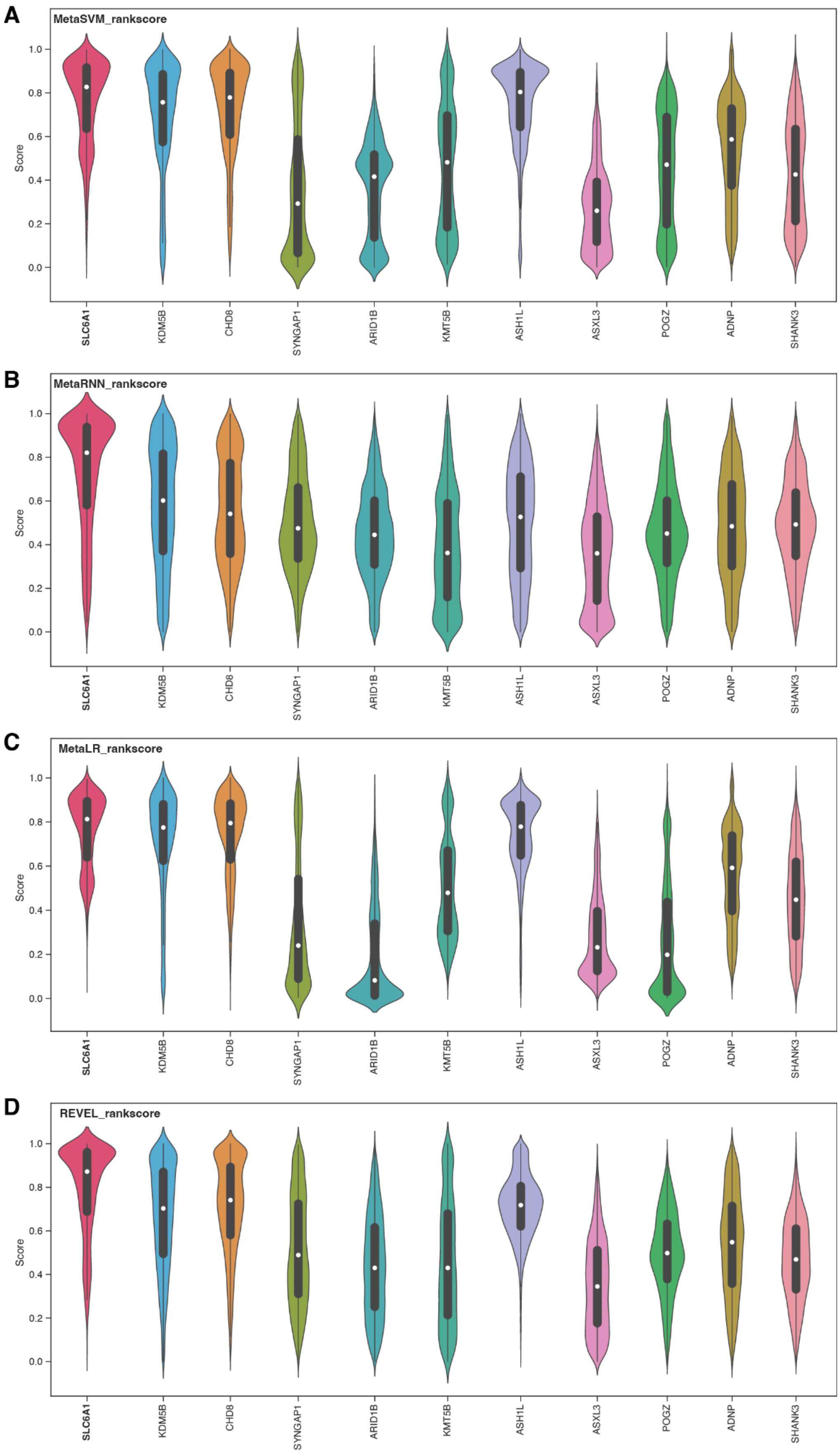
A
MetaSVM_rankscore
B
MetaRNN_rankscore
C
MetaLR_rankscore
D
REVEL_rankscore
Score
0.0
0.2
0.4
0.6
0.8
1.0
SLC6A1
KDM5B
CHD8
SYNGAP1
ARID1B
KMT5B
ASH1L
ASXL3
POGZ
ADNP
SHANK3

**Supplemental Figure 17: Distribution of missense severity scores in all possible missense variants in *SLC6A1* and ten PTV-enriched genes.** Violin plots represent the distribution of **A)** MetaSVM, **B)** MetaRNN, **C)** MetaLR, and **D)** REVEL for all possible missense variants in *SLC6A1* genes and ten equivalent PTV-enriched ASD- and NDD-associated genes (See Fig. 4B).

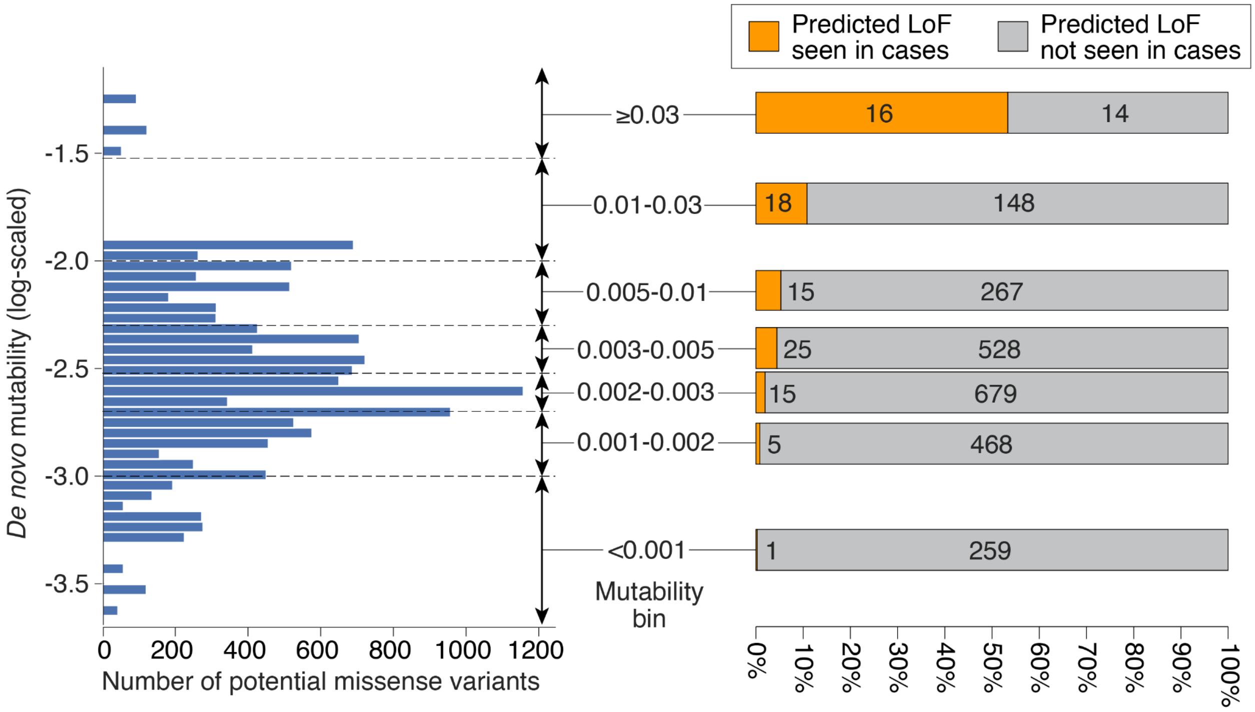


**Supplemental Figure 18. Mutability distribution of *SLC6A1* and observed frequencies for predicted loss-of-function cases.** The distribution of *de novo* mutability, estimated from base-substitution mutation rate based on triplet DNA sequence from whole-genome sequencing of families is shown across all possible missense variants in *SLC6A1* (left graph). Mutability is used to define seven bins (middle). In each of these mutability bins, the total number of missense variants predicted to be loss-of-function and observed at least once in cases is shown in orange, while the total number of missense variants predicted to be loss-of-function and not observed in cases is shown in gray. Abbreviations: LoF: Loss-of-function.
