## Supplemental methods and materials for "Haploinsufficiency underlies the neurodevelopmental consequences of *SLC6A1*/GAT-1 variants"

### Supplemental Materials and Methods

The following data generation and data processing methods are separated by the two research groups: BioMarin and UCSF. The main materials and methods section of this manuscript has a summary of combined methods from both groups.

#### Initial variant selection (BioMarin)

We identified individuals with potentially pathogenic *SLC6A1* variants from multiple sources[4,10,11,19,20,26–33] (**Table S1**). The individuals and organizations who collected and generated this original data bear no responsibility for further analysis or interpretation in this publication. After taking into account individuals represented in more than one cohort, the total number of carriers was 213. After taking into account variants present in multiple individuals and after excluding large insertion or deletion variants (indels) and most noncoding variants, we arrived at 182 variants we successfully tested (**Table S2**). Of those, 90 were variants seen in epilepsy and developmental delay patients, 11 were seen in schizophrenia patients (3 of those with a high probability of being pathogenic), 73 were variants from various sources which are seen in epilepsy cases but for which pathogenicity has not been claimed and the remaining 5 were likely benign and included as negative controls (**Table S2**).

#### Initial variant selection (UCSF)

Individuals with variants in *SLC6A1* were identified from multiple sources, including: ClinVar[18], gnomAD[19], multiple cohort studies[1–4,20], case series and reports[20], and three previously undescribed cases from the Moller group in Denmark (**Table S1**). The gnomAD aggregation database was used to select the 10 missense variants with the highest population frequencies as controls. Another 10 variants were selected as a representative group of protein truncating variants. The remaining 80 missense variants were selected based on: recurrence across multiple individuals, variants in individuals with detailed phenotyping data, and distribution across the GAT-1 protein. The full list of variants and extensive information on each can be found in **Table S2**.

#### Creation of *SLC6A1* Constructs (BioMarin)

The *SLC6A1* consensus coding-sequence (CCDS: CCDS2603.1) was synthesized by Genewiz (South Plainfield, NJ) and cloned into the pUC57- Kan plasmid with a CMV promoter driving the production of *SLC6A1*, opposite a reverse-oriented EF1-alpha promoter driving a Beta-lactamase reporter. Genscript (Piscataway, NJ) performed site-specific mutagenesis to generate 183 distinct variants. Mutagenesis was confirmed by Sanger sequencing.

#### Creation of *SLC6A1* Construct and GFP-tagged plasmids (UCSF)

The vector pCMV6-Entry containing the human cDNA of SLC6A1 (NM_003042) was purchased from OriGene (OriGene Technologies Inc., Rockville, MD, USA). The plasmid pCDNA5/FRT

SLC6A1 (WT-SLC6A1) was created by digesting the pCMV-Entry SLC6A1 (OriGene) with KpnI and NotI (NEB) to excise the SLC6A1 transcript. The mammalian expression vector pcDNA5/FRT was also digested by KpnI and NotI to create compatible restriction sites. Digested DNA sequences and size was confirmed by MCLAB sequencing (South San Francisco, CA, USA). Ligation of the digested backbone and excised cDNA were performed using T4 Ligase and instructions were followed by manufacture protocol (New England BioLabs, Inc.). The resulting reaction was transformed and the plasmid was sequenced by MCLAB, confirming proper ligation of cDNA SLC6A1 into pcDNA5/FRT. The final WT-SLC6A1 plasmid was then tested by [3H]-GABA transport assay and the reuptake of GABA was compared to the original plasmid pCMV-Entry SLC6A1. Compared to empty vector plasmid, both constructs demonstrated a 13-fold uptake of GABA, further confirming successful ligation.

The WT-SLC6A1 construct was sent to Genscript (Piscataway, NJ) and site-specific mutagenesis was performed to make 100 individual plasmids with its respective mutation. Genscript internally confirmed the quality and sequence of all plasmids. For the GFP tagged plasmids, the 249 amino acid superfolder GFP (sfGFP, ASSN: ASL68970) was synthesized by Genscript and then cloned into the c-terminal end of SLC6A1 transcript in WT-SLC6A1 construct with no spacer or linker by the same company. The final plasmid, WT-SLC6A1-sfGFP, was verified by Genscript and tested in uptake assay to compare relative uptake to WT-SLC6A1. To create the 86 GFP tagged variant plasmids, Genscript performed site-directed mutagenesis on the WT-SLC6A1-sfGFP plasmid and verified every individual variant. All final sequences are reported in **Table S3**.

#### CRISPR/Cas9 knockout of *SLC6A1* in HEK293T cells (BioMarin)

We generated a *SLC6A1* deficient cell line, using HEK293T transfected with CRISPR/Cas9 RNP using the following crRNAs targeting exon 5 with PAM in bold:

Hs.Cas9.SLC6A1.1.AD: 5’-[AGTGGCCAGCGGATCTGACC**TGG**]-3’

Hs.Cas9.SLC6A1.1.AQ: 5’-[ATACACAAGGATCCAGGCGA**TGG**]-3’

Briefly, we annealed equimolar ratios of ATTO 550 labeled tracrRNA (IDT Cat. 1075928) and the above crRNA. Twelve picomoles of the resulting gRNA were combined with 104 pmol HiFi Cas9 (IDT 1081061) to form RNPs, according to the manufacturer’s protocol. 2E5 HEK293T cells were transfected with RNP using a 96-well Shuttle nucleofector (Lonza) with Amaxa SF solution (Lonza) and program CM-138. Transfected cells were seeded into a 96-well plate and cultured for 3 days at 37°C, with 5% CO2, in 200 L of high-glucose DMEM (Invitrogen, Cat.11995-065) supplemented with 10% (v/v) Fetal Bovine Serum (VWR, Cat. 97068-085) and 1X GlutaMAX-ITM (GIBCO, Cat. 35050-061).

Next, single cells were sorted into 96-well plates containing 200 µL media using a FACSMelody cell sorter (BD Biosciences). Clones were monitored by imaging (Cell Metric, Solentim) until confluent, then expanded into six-well plates and genotyped. Genomic DNA was extracted using Quanta Extracta solution (Quanta Biosciences, Cat. 95091). DNA amplicons containing the gRNA target-region were produced using the following primers (*SLC6A1*-binding sequences in bold):

FW: 5’-<u>TCGTCGGCAGCGTCAGATGTGTATAAGAGACAG</u>**GCTCCCACCAGCTCTGTGTA**-3’

REV: 5’-<u>GTCTCGTGGGCTCGGAGATGTGTATAAGAGACAG</u>**CTATCCAGTGCCCTCCTGTCC**-3’

A second round of PCR incorporated NGS barcodes and Illumina sequencing-adapters via NexteraTM–Compatible Indexing Primers, which annealed to the adapters (underlined above) added during the first PCR. Amplicons were purified and pooled according to the manufacturer’s instructions (Illumina Document # 15031942 v05), then paired-end sequenced (149x149) on a MiSeq 550 with a v2 reagent kit (Illumina, Cat. MS-102-2002). Indels were quantified using CRISPResso (version 2.0.27) to identify a clonal population with homozygous *SLC6A1* deficiency. The clonal cell line selected for experimentation was homozygous for a 37 bp deletion in exon 5:

5’-[GCGCAACATGCATCAGATGACGGACGGGCTGGATAAGCCAGGTCCTGGATCCTTGTGTAT
TTCTGTATCTGGAAGGGTGTTGGCTGGACTGGAAAG]-3’

Resulting in the following truncated *SLC6A1* protein coding sequence:

MATNGSKVADGQISTEVSTEVSEAPVANDKPKTLVVKVQKKAADLPDRDTWKGRFDFLMSCVGY
AIGLGNVWRFPYLCGKNGGGAFLIPYFLTLIFAGVPLFLLECSLGQYTSIGGLGVWKLAPMFKGVG
LAAAVLSFWLNIYYIVIISWAIYYLYNSFTTTLPWKQCDNPWNTDRCFSNYSMVNTTNMTSAVVEF
WERNMHQMTDGLDKPGPGSLCISVSGRVLAGLERWSSTFQPHTPTSC*

The GABA uptake assay was used to confirm loss of SLC6A1 activity in the knockout line.

#### Cell culture preparation of HEK293-FlpIn cells and creation of transient and stable cell lines (UCSF)

Human embryonic kidney cells (HEK293) with the Flp-In integration system (Life Technologies Corporation, Carlsbad, CA, USA) were used for this study. Cell culture preparation of HEK293 cells requires growth and maintenance in Dulbecco’s Modified Eagle Medium (DMEM) media with 100 units/mL penicillin, 100 units/mL streptomycin, and 10% fetal bovine serum (FBS). Stable or transient cell lines were transfected with the WT-SLC6A1 plasmid or the empty vector pcDNA5/FRT (EV) cDNA in HEK293 cells. The reagent Lipofectamine LTX (Life Technologies Corporation, Carlsbad, CA, USA) was used to transfect cells transiently in triplicates on 3 separate days or stably. Appropriate DNA to LTX ratios were followed according to the manufacturer's protocol (final 2:1). Using poly-D-lysine treated 96-well plates and an optimized seeding density of $3.3x10^5$ cells/well, cells were reverse transfected with 200 ng of DNA and 0.4 µL of Lipofectamine LTX in Opti-MEM medium (Life Technologies Corporation) to transfect each well. FBS only DMEM media is used on seeding and transfecting day. Plates were left for 45 minutes in the hood at room temperature before placing in the incubator with 5% $CO_2$ at a stable 37 °C to allow cells to settle and spread out evenly on the well surface. Cells were grown for 48 hours post-reverse transfection then assayed for GABA transport. To create stable cell lines,

HEK293 Flp-In cells were transfected using Lipofectamine LTX with a 1:2 DNA (in pcDNA5/FRT mammalian expression vector) to LTX ratio and a 1:9 ratio of DNA to pOG44 (Flp-Recombinase expression vector) in a 6-well plate treated with poly-D-lysine. After 48 hours, cells were split into a 10 mm dish and allowed to sit overnight. Positive clone selection began the next day by using fresh DMEM media with 100 µg/mL of Hygromycin B (Life technologies Corporation) and changed every 2-3 days for about two weeks.

#### Transfection of Variants and GABA uptake assay (BioMarin)

HEK293T *SLC6A1*-deficient cells were transfected in sextuplicate, using a 96-well Shuttle nucleofector (Lonza) integrated onto a MICROLAB STAR liquid-handling robot (Hamilton Robotics, Reno, NV). Briefly, 250 ng of plasmid per 300,000 cells was transfected using Amaxa solution SF (Lonza) and program CM-138. Cells were seeded onto 96-well plates, then cultured for 3 days in 200 µL media, as described above.

Prior to the reuptake assay, cells were centrifuged at 300 xg for 5 min and all media was aspirated. Cells were then incubated at 37°C for 15 min with 80 µL of Pre-Incubation solution containing 140 mM NaCl, 5 mM KCl, 2 mM CaCl2, 1 mM MgSO4, 2 nM Glucose, 2.5 mM HEPES at pH 7.4 and osmolarity 310. Pre-incubation solution was aspirated, following centrifugation at 300 xg for 5 min. Cells were incubated for 30 min at room temperature with 100 µL of 4800 nM d6-GABA (SIGMA, Cat. 615587) in a pre-incubation solution. After centrifuging for 5 min at 300 xg, the uptake solution was aspirated, and cells were washed twice with 80 µL preincubation solution. Supernatant was aspirated and plates were flash frozen at -80°C. Cells were harvested by adding 60 µL MPER (Thermo Fisher Scientific, Cat. 78501) and lysed with rigorous pipetting. 10 µL of lysate was used to determine protein concentrations with a Pierce BCA Protein Assay (Thermo Fisher Scientific, Cat. 23227), according to the manufacturer's instructions.

Beta-lactamase (BLA) activity was determined by measuring hydrolysis of d7-penicillin G (Toronto Research Chemicals, Cat. B288600) into d7-5R,6R-benzylpenicilloic acid (d7-BPA). The BLA Assay Cocktail consisted of 60 µM d7-penicillin G substrate and 1.5 µM 5R,6R-benzylpenicilloic acid (Toronto Research Chemicals, Cat. B288593), in 50 mM Tris-HCl (pH 7.5±0.02). For absolute quantification, separate BPA and d6-GABA standard curves were prepared with the following ranges: 120 µM, 60 µM, 20 µM, 10 µM, 2 µM, 1 µM and 200 nM for BPA; and 6 µM, 3 µM, 1 µM, 500 nM, 100 nM, 50 nM, 10 nM, 1 nM and 500 pM for GABA. To avoid hydrolysis, d7-penicillin G and BPA were stored dry, under inert gas at -20°C, and the BLA Assay Cocktail and BPA standards were prepared fresh. BPA standards and lysates were processed in parallel with sample lysates. Briefly, 20 µL of BLA Assay Cocktail was combined with 20 µL lysate or BP A standard, then incubated for 1 hour at 37°C, on an orbital shaker set to 500 rpm. The reaction was quenched with 160 µL of acetonitrile containing 25nM d2-GABA (Sigma, Cat. 617458), then vortexed at 1500 rpm for 5 min and centrifuged at 3000×G for 10 min. 100 µL of supernatant was combined with an equal volume of HPLC-grade water and reserved for the BLA-assay injection. An additional 20 µL of supernatant was combined with 10 µL 100 mM Sodium Carbonate in a 96-well plate, then vortexed for 5 min at 1500 rpm. GABA was derivatized with 10 µL 6% (v/v) Benzoyl-Chloride in acetonitrile, vortexing at 1500 rpm for 5 min. The reaction was quenched with 0.2% Formic Acid (v/v) in water, then centrifuged at 4000 rpm for 4 min, to produce the final GABA analyte.

UPLC-MS/MS analysis was performed on an Agilent 6495 Triple-Quadrupole LC/MS, equipped with an Agilent 1290 Infinity II series HPLC. Separation of analytes was achieved using an Acquity UPLC BEH C18 1.7 µm, 2. x 50 mm column (Waters Crop., Cat. 176001692) at 40°C. Mobile phase A (MPA) contained water with 0.1% formic acid, and mobile phase B (MPB) was acetonitrile with 0.1% formic acid. The BLA and GABA analytes were analyzed in separate, 10 µL injections, utilizing the same LC-MSMS instruments and mobile phases, but different gradients. For the BLA assay, a flowrate of 0.4 mL/min was applied with the following gradient: 0-1 min, 5% mobile phase B; 1-2 min, 5%-95% MPB; 2-2.9 min, 95% MPB; 2.9-3 min 5% MPB. The enzymatic product (d7-5R,6R-benzylpenicilloic acid) and internal standard (5R,6R-benzylpenicilloic acid) were detected through multiple reaction monitoring (MRM) using the following transitions: 360.3 > 160 and 353.3 > 160, respectively. For the GABA assay, the flowrate was set to 0.4 mL/min with the following gradient: 0-4 min, 5-42% mobile phase B; 4-4.4 min, 42%-95% MPB; 4.4-4.9 min 95% MPB; 4.9-5% MPB. We monitored the GABA transported into cells by SLC6A1 (d6-GABA-BZ, following derivatization) and the internal standard (benzoylated d2-GABA) by MRM, using distinct transitions: d6-GABA-BZ, 214.1 > 105; d2-GABA-BZ, 210.1 > 105. In both assays, the dwell time was set to 50 msec. The cone voltage in both assays was 4500 V and the collision energy varied, using 17 V in the GABA assay and 11 V for the BLA method. Method scheduling was performed in Mass Hunter (Agilent, version 10.0). Peak calling and standard-curve calculations were executed in QQQ Quantitative Analysis (Agilent, version 10.1).

**[$^3$H]-GABA Transport Assay (UCSF)**

HEK293 cells were transiently transfected as described above in 96-well plates. Uptake of [$^3$H]-GABA was assayed 48 hours post-reverse transient transfection or 24 hours after plating stable cells. Warm choline buffer (150 mM choline chloride, 5 mM $KP_i$, 0.5 mM $MgSO_4$, and 0.3 mM $CaCl_2$, pH 7.4) was used to wash cells twice on the day of assaying. Cells were then incubated with the same choline solution for 30 minutes in the incubator. Transport solution containing 150 mM NaCl, 5 mM $KP_i$, 0.5 mM $MgSO_4$, and 0.3 mM $CaCl_2$ at pH 7.4 was used to dilute 4 µCi of [$^3$H]-GABA (35 Ci/mmol). The choline solution from each well was then replaced with prepared [$^3$H]-GABA transport solution for an incubation of 10 minutes. Prior to beginning this study, the transport assay was studied at an incubation of 10 minutes, 15 minutes and 20 minutes. No difference in uptake was apparent. After 10 min, the reaction was then slowed down and terminated by washing the cells twice with ice-cold NaCl transport buffer only. Cells were completely aspirated of solution and lysed with 0.28 mL of lysis buffer (0.1N sodium hydroxide (NaOH) and 1% v/v SDS solution). After shaking the cells with lysis buffer for 60-90 min, 0.25 mL of cell lysate was transferred to scintillation fluid (EcoLite(+)™, MP Biomedicals, Santa Ana, CA, USA) for scintillation counting on the Beckman Coulter LS6500. The remaining lysate (20 µL) was used to measure total protein using the BCA assay kit. Uptake values were corrected for protein concentration using BCA values for each well.

#### Transfections for dominant negative effect (UCSF)

Dominant negative study design was optimized from previous groups[56–58]. HEK293 cells were seeded at an optimized cell density of $3.3x10^5$ cells/well in a poly-D-lysine treated 96-well plate and reverse transfected with respective plasmid(s) on the same day. The reagent Lipofectamine LTX (Life Technologies Corporation, Carlsbad, CA, USA) was used to transfect cells transiently in triplicates. Using the manufacturing protocol, we optimized our DNA to LTX ratio to 2:1, or 2 µL of Lipofectamine LTX for every 1 µg of DNA. A master mix with DNA, LTX and Opti-MEM medium (Life Technologies Corporation) is prepared according to the manufacturer's protocol of the LTX reagent. For dominant negative studies, we tested a 1:1 ratio of two distinct plasmids co-transfected in the same well, with a total of 100 ng per well. For example, the WT-SLC6A1 plasmid was mixed with a mutated SLC6A1 plasmid (50 ng WT:50 ng Mut) during master mix preparation. For those wells transfected with control or mutant only, we tested 200 ng, 100 ng, and 50 ng of the plasmid alone to compare activity to co-transfections and evaluate transfection efficiency. FBS only DMEM media is used on seeding/transfecting day. Plates were left for 45 minutes in the hood at room temperature before placing in the incubator with 5% $CO_2$ at a stable 37 °C to allow cells to settle and spread out evenly on the well surface. Cells were grown for 48 hours post-reverse transfection then assayed for GABA transport as described above (UCSF).

#### Data analysis (BioMarin)

BLA activity (nmol/hr/mg_protein) was determined using nanomoles d7-BPA, as calculated from the BPA standard curve, then divided by the incubation time (1 hour). This rate was further divided by the milligrams of lysate protein, as a proxy for variable cell counts between wells. SLC6A1 re-uptake activity (nmol/hr/mg_protein) was calculated similarly, by determining the nanomoles of d6-GABA from the d2-GABA standard curve, then dividing by both the incubation time (0.5 hours) and milligrams of protein in the lysate. To account for variable transfection and expression efficiencies, SLC6A1 re-uptake activity (nmol/hr/mg_protein) was then divided the BLA activity, to derive the GABA uptake normalized to BLA activity. While the BLA-normalized GABA activity is an accurate measure of a variant's transport activity, the functional consequence of a mutation is best described in relation to wildtype-GAT-1 transport—to convey this, we calculated "Percent Wildtype" activities, where each variant's BLA-normalized SLC6A1-activity was divided by the average BLA-normalized SLC6A1 activity of all wildtype replicates present within an assay plate reported in **Table S4**. Figures were generated in Python matplotlib, seaborn, or SankeyMATIC (https://sankeymatic.com/). Figure 6 was generated using Protter (version 1.0)[59].

#### Distribution of measurement variation (BioMarin)

To account for variability in transfection efficiencies, GABA reuptake activity was normalized to activity of a Beta-lactamase (BLA) reporter present on the *SLC6A1* expression construct. BLA activity was determined by measuring hydrolysis of d7-penicillin G. While the BLA-normalized GABA reuptake activity is an accurate measure of a variant's transport activity, there exists experimental variability that is biased towards *SLC6A1* variants with high activity. The experimental variation is heightened due to the high sensitivity of the mass spectrometry assays which can detect even slight variations that could be caused by miniscule pipetting errors. Ultimately biological replicates were included to assess the variability in transporter activity of SLC6A1 variants which was normalized to activity in wildtype cells.

#### Stepwise linear regression

To build a predictor of missense severity in *SLC6A1*, we annotated all 179 missense variants with functional data from BioMarin and/or UCSF against estimates of mutability from whole-genome sequence of families[60], numerous predictors of missense severity using annoVar protocol ‘dbnsfp42a’ and build ‘hg38’[[21]], and estimates of protein stability: DynaMut[61] and DeepDDG[62]. Linear regression was performed for all 81 quantitative variables against 147 missense variants with values for all variables (**Table S2**). The lowest p-value was achieved by ClinPred_rankscore ($P=2.6 \times 10^{-23}$, $R^2=0.49$). Linear regression was repeated for the other 80 variables, but ClinPred_rankscore was included in the model too. This process of picking the top ranked variable by P-value was repeated for 10 rounds, achieving a model with an $R^2$ of 0.63. Three variables were only available for a subset of missense variants: MutPred (166 variants), DynaMut (160 variants), DeepDDG (160 variants). Of these only MutPred_score was included in the model (step 4, P=0.02). To assess if increasing the number of variants improved the model, we repeated the stepwise linear regression excluding MutPred, DynaMut, and DeepDDG. This yielded 177 missense variants with functional data and annotations for all 78 quantitative metrics. Repeating the linear regression (**Table S6**), the lowest p-value was again achieved by ClinPred_rankscore ($P=7.1 \times 10^{-35}$, $R^2=0.58$, beta=-1.085, intercept=0.204) and ten rounds of stepwise linear regression led to a model with an $R^2$ of 0.68. Given the risk of overfitting to a relatively small dataset, we elected to use the ClinPred_rankscore metric alone (scaled from no predicted impact at 0 to severe predicted impact at 1).

To extrapolate the GABA uptake functional data to all possible missense variants in *SLC6A1* we used the ENST00000287766.10 transcript for the ENSG00000157103.12 *SLC6A1* gene as defined by GENCODE v39 and the GRCh38/hg38 genome build to define every possible DNA variant in *SLC6A1* (**Table S7**). Variants were annotated as for the variants with GABA uptake data and the ClinPred_rankscore was used to predict GABA uptake (**Table S7**).

#### Random forest analysis

We performed a random forest model to provide a second regression method predicting functional uptake. We used the R packages ‘randomForest’ and ‘caret’, and R defaults for number of trees sampled, 500, and number of independent variables sampled at each split, 2. Built in out-of-bag error estimates were used to evaluate performance.

Initial independent variables included location of mutation (represented by exon number), initial amino acid, altered amino acid, and mutability score. Using the training set consisting of 179 missense variants with protein uptake set as the outcome variable, we analyzed 75 pre-existing prediction tools to optimize variance explained by the model. We iteratively added the best performing tool until addition of another tool led to <1% improvement in variance explained. MetaRNN raw score, ClinPred Rankscore, and Provean converted rankscores were added to the model in that order via this method. This model resulted in between 54.13-55.18% of the variance explained over the course of ten runs.

This model was then applied to known possible missense variants of the GAT-1 protein. Predicted uptake for all possible missense variants are included in **Table S7**. When converting continuous uptake outcomes to equivalent categorical outcomes prespecified in main body of the paper, 414 fell into the Severe LoF range, 1,735 were predicted to fall into the LoF range, 1,803 were predicted to be wild type, and 0 were predicted in the gain of function range.
